# Community factors and excess mortality in the COVID-19 pandemic in England, Italy and Sweden

**DOI:** 10.1101/2022.04.26.22274332

**Authors:** Brandon Parkes, Massimo Stafoggia, Daniela Fecht, Bethan Davies, Carl Bonander, Francesca de’ Donato, Paola Michelozzi, Frédéric B. Piel, Ulf Strömberg, Marta Blangiardo

**Affiliations:** UK Small Area Health Statistics Unit, Department of Epidemiology and Biostatistics, School of Public Health, Imperial College London, Norfolk Place, London, UK; 2MRC Centre for Environment and Health, Department of Epidemiology and Biostatistics, School of Public Health, Imperial College London, Norfolk Place, London, UK; Department of Epidemiology, Lazio Regional Health Service, ASL Roma 1, Rome, Italy; National Institute for Health Research Health Protection Research Unit in Chemical and Radiation Threats and Hazards, Department of Epidemiology and Biostatistics, School of Public Health, Imperial College London, Norfolk Place, London, UK; Health Economics and Policy, School of Public Health and Community Medicine, Institute of Medicine, University of Gothenburg, Box 463, 405 30 Gothenburg, Sweden; National Institute for Health Research Health Protection Research Unit (NIHR HPRU) in Health Impact of Environmental Hazards, Imperial College London, UK; School of Public Health and Community Medicine, Institute of Medicine, Sahlgrenska Academy at University of Gothenburg, Sweden; Department of Research and Development, Region Halland, Halmstad, Sweden

**Keywords:** SARS-CoV-2, spatial disease mapping, small-area, Bayesian smoothing, community factors

## Abstract

**Background:** Analyses of COVID-19 suggest specific risk factors make communities more or less vulnerable to pandemic related deaths within countries. What is unclear is whether the characteristics affecting vulnerability of small communities within countries produce similar patterns of excess mortality across countries with different demographics and public health responses to the pandemic. Our aim is to quantify community-level variations in excess mortality within England, Italy and Sweden and identify how such spatial variability was driven by community-level characteristics.

**Methods:** We applied a two-stage Bayesian model to quantify inequalities in excess mortality in people aged 40 years and older at the community level in England, Italy and Sweden during the first year of the pandemic (March 2020–February 2021). We used community characteristics measuring deprivation, air pollution, living conditions, population density and movement of people as covariates to quantify their associations with excess mortality.

**Results:** We found just under half of communities in England (48.1%) and Italy (45.8%) had an excess mortality of over 300 per 100,000 males over the age of 40, while for Sweden that covered 23.1% of communities. We showed that deprivation is a strong predictor of excess mortality across the three countries, and communities with high levels of overcrowding were associated with higher excess mortality in England and Sweden.

**Conclusion:** These results highlight some international similarities in factors affecting mortality that will help policy makers target public health measures to increase resilience to the mortality impacts of this and future pandemics.

## Introduction

The Coronavirus Disease 19 (COVID-19) pandemic has been spreading throughout Europe since early 2020 causing an estimated 580,000 deaths by the end of 2020.^1^ The impact of the pandemic has varied widely across Europe resulting in speculation as to the possible reasons for higher mortality in some countries. The multitude of factors affecting mortality and the heterogeneity of data collected in different countries make accurate international comparisons challenging. Existing comparison studies have examined COVID-19 mortality between countries down to the NUTS3 regional level^2-4^ but not for smaller areas that would allow the identification of the community characteristics which affect the risk of excess mortality in local populations.^5^

Here we study England, Italy and Sweden, three European countries differently affected by the pandemic and characterised by very different public health responses and policies. Italy was the first European country to be hit by large numbers of SARS-CoV-2 infections, soon followed by England and the rest of Europe. By 28 February 2021, after a year of COVID-19 in Europe, cumulative mortality attributed to COVID-19 in England, Italy and Sweden had reached 1,930, 1,618 and 1,262 deaths per million people respectively.^6, 7^

Here we aim to quantify community-level variations in excess mortality within the three countries and to identify how such spatial variability was driven by community-level characteristics. We analysed data on all-cause mortality at ages 40 years and over for 6,791, 7,900 and 5,985 community-level areas in England (Middle-layer Super Output Areas, MSOAs), Italy (municipalities) and Sweden (Demografiska statistikområden, DeSOs) respectively, from 1 March 2020 to 28 February 2021.

## Methods

### Excess mortality

Excess mortality, typically calculated as a counterfactual comparison using data from years prior to the pandemic, is an overall measure of the mortality impact of COVID-19. It includes not just deaths known to be caused by SARS-CoV-2 infection, but also deaths where SARS-CoV-2 infection was undiagnosed and deaths resulting from behavioural, social and healthcare changes that accompanied interventions to reduce SARS-CoV-2 infections.

### Study period and units of analysis

We considered mortality data over a one-year study period (1 March 2020 to 28 February 2021) and a five-year comparison period (1 March 2015 to 29 February 2020). The study period and comparison period were chosen to reflect the different dynamics of the pandemic in England, Italy and Sweden and to minimise the effect of the vaccination rollout which commenced on 8 December 2020 in England and 27 December in Italy and Sweden.^8-10^ Due to the low numbers of deaths in people under the age of 40 years^2, 11^, we limited the analysis to people over that age.

To investigate the association of community characteristics with excess mortality we included data at small area level with a view to maintaining consistency across England, Italy and Sweden. For England, we used MSOAs (mean population: 8,288, range: 2,224 to 26,513). In Sweden, we used DeSOs, (mean population: 1,726,range 663 – 5,291).^12^ Both MSOAs and DeSOs have been defined based on population size. For Italy, the only comparable geographical units for which data on mortality and covariates were available was the municipality which has a much broader population range (mean: 7,548, range: 30 – 2,808,293).^13^

### Characteristics of Communities

We considered community characteristics measuring deprivation, air pollution, living conditions, population density and movement of people. These were used as covariates in our model to quantify their associations with excess mortality. A summary of community characteristics is presented in Table 1. Covariates were provided as five categories (overcrowding and deprivation for Italy) or continuous data which we divided into quintiles, giving, in each category, ∼1,358 MSOAs (England), ∼1,580 municipalities (Italy) and ∼1,197 DeSOs (Sweden).

**Table 1:**
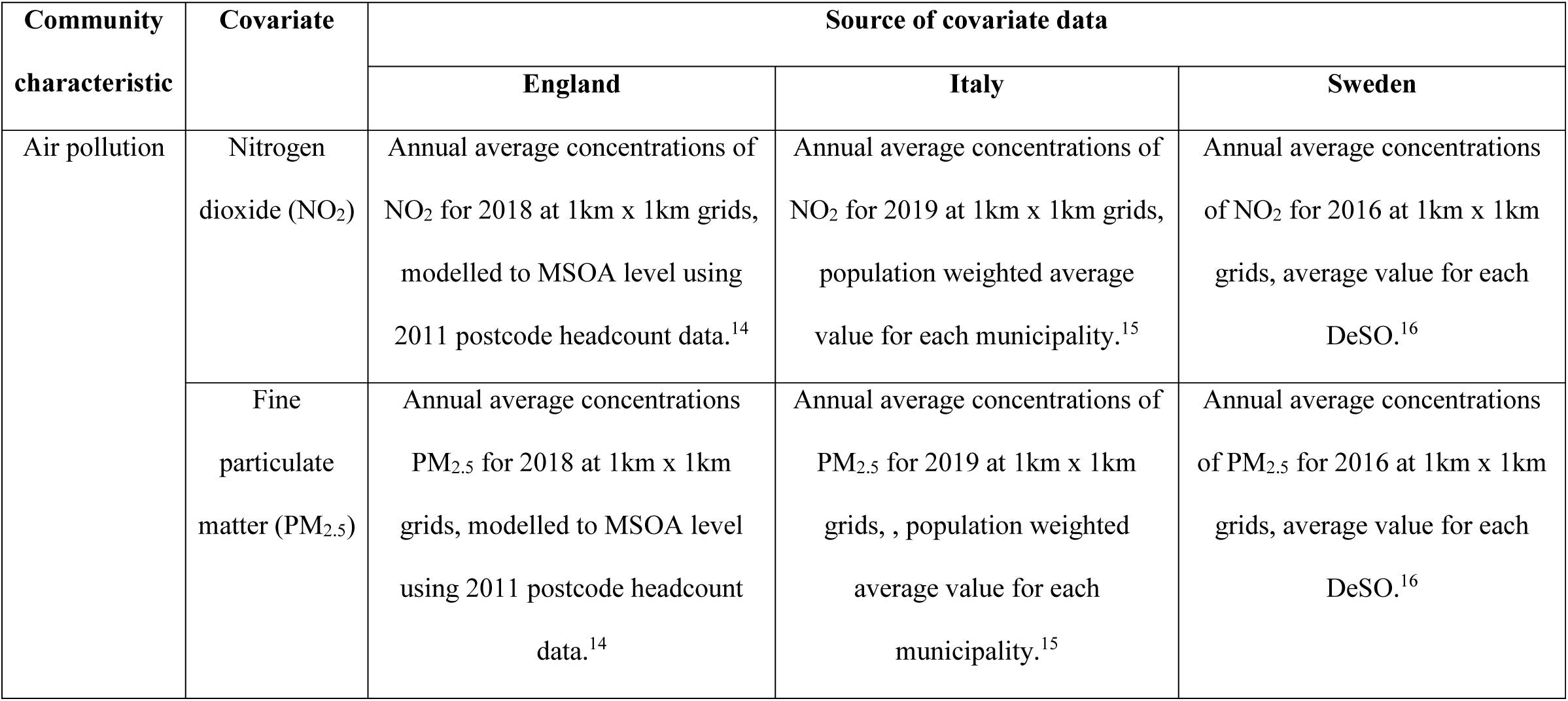
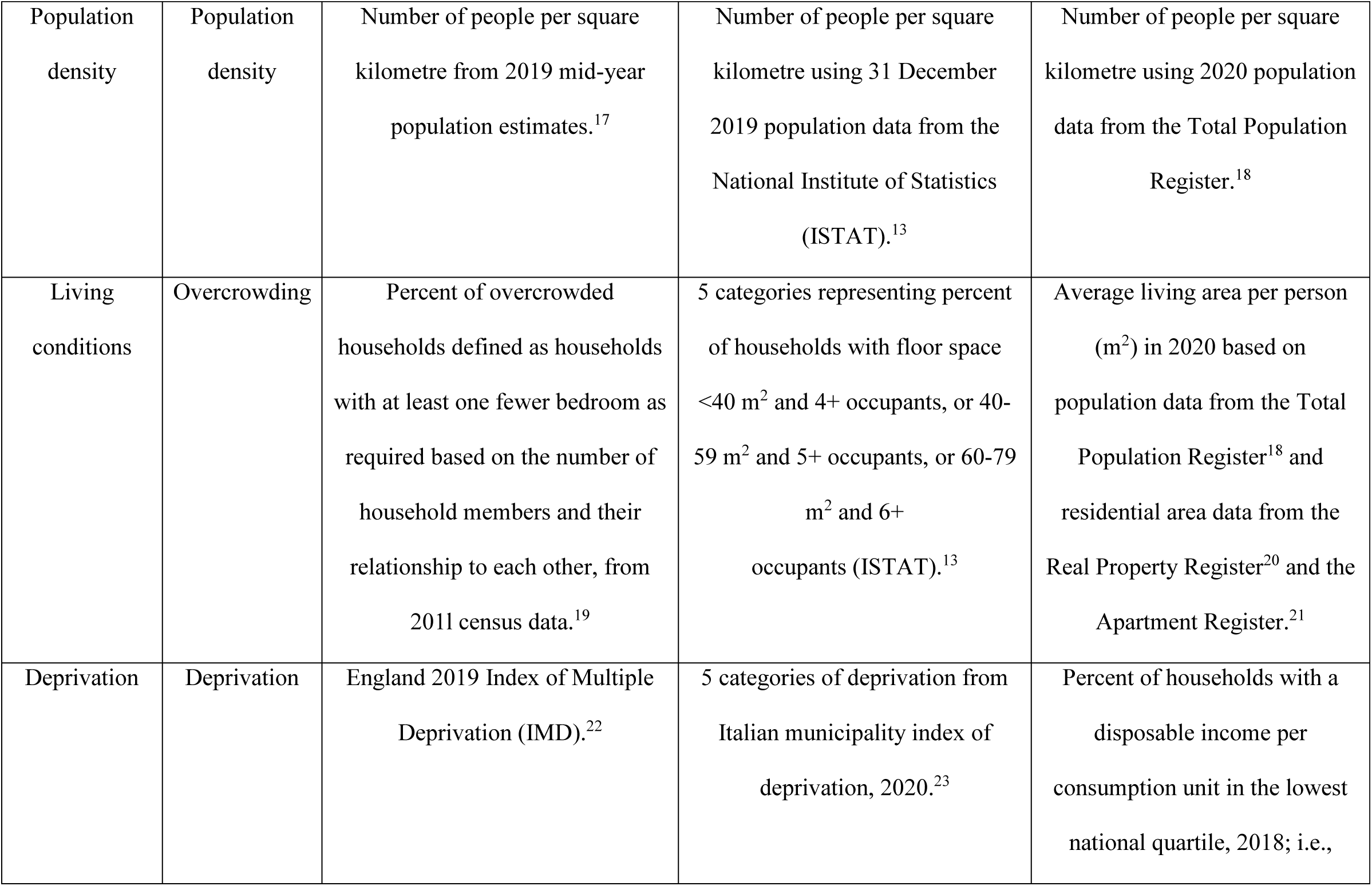
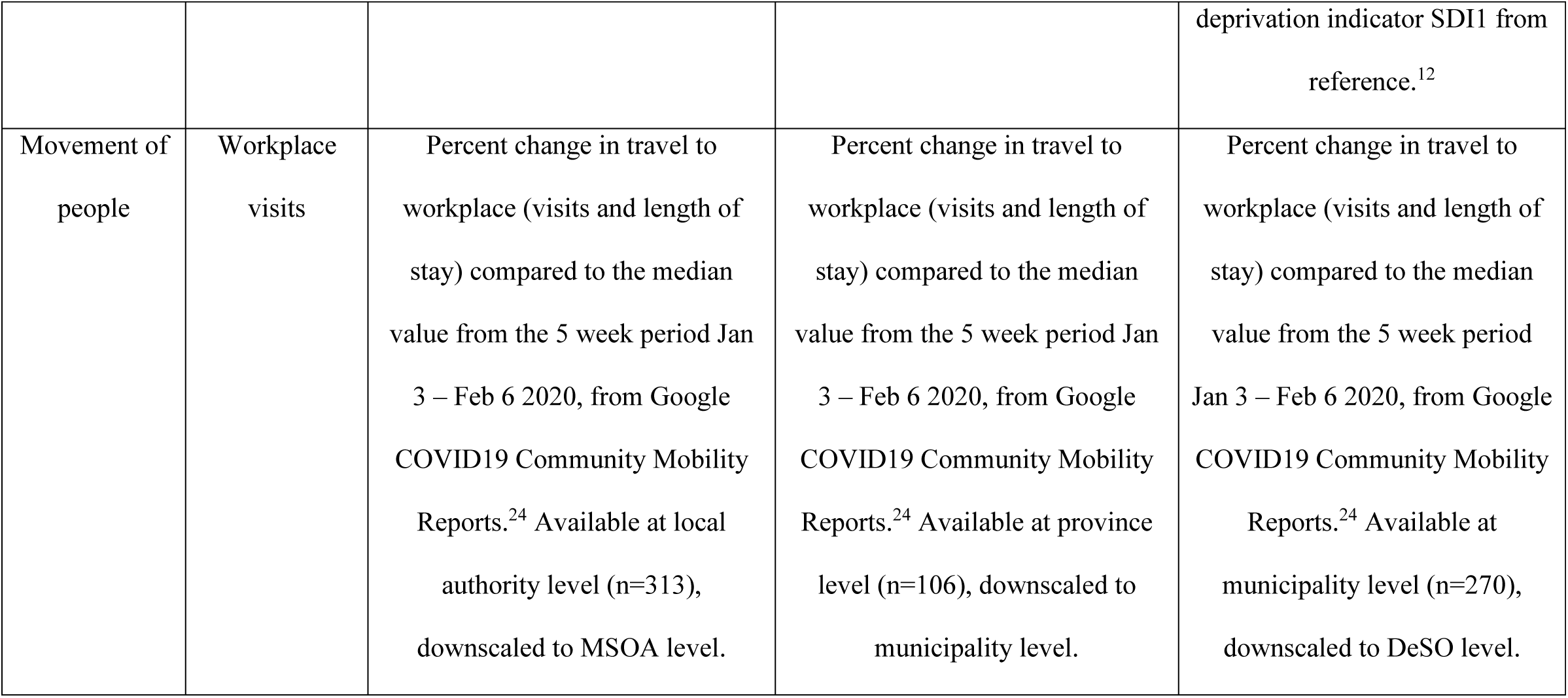
Covariate data for England, Italy and Sweden used in the analysis. Where not specified as categorical data the variables are continuous which we then divided into quintiles. Categories or quintiles were ordered based on the expected relationship between the covariate and excess mortality, with the highest category/quintile associated with higher excess mortality (e.g. high air pollution associated with higher excess deaths, but lowest change in workplace visits associated with higher excess deaths).

| Community characteristic | Covariate | Source of covariate data |  |  |
| --- | --- | --- | --- | --- |
|  |  | England | Italy | Sweden |
| Air pollution | Nitrogen dioxide (NO<sub>2</sub>) | Annual average concentrations of NO<sub>2</sub> for 2018 at 1km x 1km grids, modelled to MSOA level using 2011 postcode headcount data.<sup>14</sup> | Annual average concentrations of NO<sub>2</sub> for 2019 at 1km x 1km grids, population weighted average value for each municipality.<sup>15</sup> | Annual average concentrations of NO<sub>2</sub> for 2016 at 1km x 1km grids, average value for each DeSO.<sup>16</sup> |
|  | Fine particulate matter (PM<sub>2.5</sub>) | Annual average concentrations of PM<sub>2.5</sub> for 2018 at 1km x 1km grids, modelled to MSOA level using 2011 postcode headcount data.<sup>14</sup> | Annual average concentrations of PM<sub>2.5</sub> for 2019 at 1km x 1km grids, , population weighted average value for each municipality.<sup>15</sup> | Annual average concentrations of PM<sub>2.5</sub> for 2016 at 1km x 1km grids, average value for each DeSO.<sup>16</sup> |
| Population density | Population density | Number of people per square kilometre from 2019 mid-year population estimates.<sup>17</sup> | Number of people per square kilometre using 31 December 2019 population data from the National Institute of Statistics (ISTAT).<sup>13</sup> | Number of people per square kilometre using 2020 population data from the Total Population Register.<sup>18</sup> |
| Living conditions | Overcrowding | Percent of overcrowded households defined as households with at least one fewer bedroom as required based on the number of household members and their relationship to each other, from 2011 census data.<sup>19</sup> | 5 categories representing percent of households with floor space <40 m<sup>2</sup> and 4+ occupants, or 40-59 m<sup>2</sup> and 5+ occupants, or 60-79 m<sup>2</sup> and 6+ occupants (ISTAT).<sup>13</sup> | Average living area per person (m<sup>2</sup>) in 2020 based on population data from the Total Population Register<sup>18</sup> and residential area data from the Real Property Register<sup>20</sup> and the Apartment Register.<sup>21</sup> |
| Deprivation | Deprivation | England 2019 Index of Multiple Deprivation (IMD).<sup>22</sup> | 5 categories of deprivation from Italian municipality index of deprivation, 2020.<sup>23</sup> | Percent of households with a disposable income per consumption unit in the lowest national quartile, 2018; i.e., deprivation indicator SDI1 from reference. <sup>12</sup> |
| Movement of people | Workplace visits | Percent change in travel to workplace (visits and length of stay) compared to the median value from the 5 week period Jan 3 – Feb 6 2020, from Google COVID19 Community Mobility Reports.<sup>24</sup> Available at local authority level (n=313), downscaled to MSOA level. | Percent change in travel to workplace (visits and length of stay) compared to the median value from the 5 week period Jan 3 – Feb 6 2020, from Google COVID19 Community Mobility Reports.<sup>24</sup> Available at province level (n=106), downscaled to municipality level. | Percent change in travel to workplace (visits and length of stay) compared to the median value from the 5 week period Jan 3 – Feb 6 2020, from Google COVID19 Community Mobility Reports.<sup>24</sup> Available at municipality level (n=270), downscaled to DeSO level. |

### Data Sources

For England, mortality data were obtained from the UK Small Area Health Statistics Unit’s (SAHSU) national mortality registrations supplied by the Office for National Statistics (ONS). We used mortality data for 2015-2020 and provisional data for 2021 supplied by the ONS in July 2021 which included date of death and place of residence of the deceased. Annual population was from ONS mid-year (June) population estimates for MSOAs in England, 2015 to 2019. For 2020, the population was a combination of ONS population estimates and linear regressions designed to minimise the effect of the deaths in the first wave of the pandemic in England (March-May 2020) (Supplementary Section 1).

For Italy, mortality and population data was retrieved from the National Institute of Statistics (ISTAT) website.^25^ Population data assessed on 31 December 2015 was used for the period 1 March 2015 to 28 February 2016, and so on.

For Sweden, Statistics Sweden supplied geocoded mortality and population data to each DeSO, based on the mortality registrations and residential address data in the Total Population Register.^18^ Population data assessed on 31 December 2015 was used for the period 1 March 2015 to 29 February 2016, and so on.

### Statistical methods

This study extends previous work by SAHSU examining excess mortality in England during the first wave of the COVID-19 pandemic.^5^ We carried out all analyses for males and females and for each country separately. We split the data into four age groups: 40-59 years; 60-69 years; 70-79 years; and 80+ years.

Given the small population at MSOA and DeSO level, as well for many of the Italian municipalities, we built a two stage Bayesian spatial model, which overcomes potential issues of instability of mortality estimates to obtain estimates of excess death rates based on data for each community, its neighbouring communities and the national average. In the first stage, we specified a log linear model for death rates over the comparison period by year, spatial unit and age groups. The model included year and age specific terms, as well as a random effect which accounted for spatial correlation through a neighbourhood structure. The model outputs smoothed estimates of death rates for the comparison period.

In the second stage, we modelled death rates by spatial units and age groups as the product of the ones estimated from the first stage and an additional term which represents the excess. On the log-transformed excess, we specified a linear model where we included an age specific term, a spatial random effect and the categories of the covariates. More details are provided in Supplementary Section 2.

Uncertainty was considered in each stage by drawing 100 samples from the posterior distribution of each parameter, giving a total of 10,000 samples from which we defined 95% credible intervals (CI, 2.5^th^ to 97.5^th^ percentiles). The numbers of samples ensures accuracy of the credible interval estimates, while limiting computational burden as discussed in.^5^ We fitted the models using the Integrated Nested Laplace Approximation (INLA), through the R-INLA software package (http://www.r-inla.org/).^26, 27^

To show the change in death rates associated with the covariate categories, we report posterior mean and 95% CIs of mortality rate ratios. We calculated excess deaths per 100,000 people for each area from the deaths predicted in the study period using the parameters learnt from the comparison period (which represent the estimated mortality under the alternative scenario of the absence of the pandemic) subtracted from the deaths registered in the study period (see Supplementary Section 2 for more details). We map the posterior mean excess mortality and the posterior probability of an excess above zero.

As sensitivity analyses, we ran models: i) without accounting for local spatial correlation; ii) using continuous variables instead of quintiles for the covariates which were directly comparable across the three countries (NO_2_, PM_2.5_ and population density)

## Results

### Spatial patterns of excess mortality

During the study period, 583,255, 751,893 and 97,997 people died in England, Italy and Sweden respectively compared to an average of 483,447, 635,168 and 88,709 deaths per year from the comparison period (1 March 2015 to 29 February 2020). Table 2 gives a summary of the distribution of excess deaths across the community-level areas. England and Italy showed positive excess mortality in more than 90% of areas, while for Sweden values were lower at 71% and 55% for males and females respectively. When considering >300 excess deaths per 100,000 population over 40, almost half of areas matched this criterion for males in England and Italy, but only 23% of areas did in Sweden.

**Table 2.** Summary of results of excess mortality for England, Italy and Sweden

|  | England<br>(n = 6,791 MSOAs) |  | Italy<br>(n = 7,900<br>Municipalities) |  | Sweden<br>(n = 5,985 DeSOs) |  |
| --- | --- | --- | --- | --- | --- | --- |
|  | Male | Female | Male | Female | Male | Female |
| Areas with: |  |  |  |  |  |  |
| excess mortality | 6,520<br>(96.0%) | 6,253<br>(92.1%) | 7,399<br>(93.7%) | 7,262<br>(91.9%) | 4,058<br>(71.4%) | 3,317<br>(55.4%) |
| >100 excess deaths per<br>100,000 population over 40 | 5,923<br>(87.2%) | 5,128<br>(75.5%) | 6,304<br>(79.8%) | 6,041<br>(76.5%) | 2,934<br>(49.0%) | 2,088<br>(34.9%) |
| > 200 excess deaths per<br>100,000 population over 40 | 4,675<br>(68.8%) | 3,466<br>(51.0%) | 4,912<br>(62.2%) | 4,626<br>(58.6%) | 2,019<br>(33.7%) | 1,374<br>(23.0%) |
| > 300 excess deaths per<br>100,000 population over 40 | 3,266<br>(48.1%) | 2,011<br>(29.6%) | 3,621<br>(45.8%) | 3,204<br>(40.6%) | 1,381<br>(23.1%) | 993<br>(16.6%) |
| posterior probability of<br>excess deaths at least 90% | 4,246<br>(62.5%) | 3,001<br>(44.2%) | 4,576<br>(57.9%) | 4,165<br>(52.7%) | 532<br>(8.89%) | 338<br>(5.65%) |
| Posterior probability of<br>deficit deaths at least 90% | 5<br>(0.07%) | 15<br>(0.22%) | 8<br>(0.10%) | 14<br>(0.17%) | 29<br>(0.49%) | 54<br>(0.90%) |
| Percent of population over 40 living in areas with: |  |  |  |  |  |  |
| excess mortality | 95.7% | 91.2% | 91.7% | 90.3% | 66.3% | 57.1% |
| > 100 excess deaths per<br>100,000 population over 40 | 86.1% | 73.7% | 78.0% | 66.8% | 47.7% | 35.8% |
| > 200 excess deaths per<br>100,000 population over 40 | 66.7% | 49.5% | 52.3% | 48.0% | 32.8% | 24.1% |
| > 300 excess deaths per<br>100,000 population over 40 | 45.7% | 28.7% | 35.4% | 29.9% | 22.3% | 17.6% |
| posterior probability of<br>excess deaths at least 90% | 60.6% | 42.7% | 64.9% | 56.4% | 8.78% | 6.07% |

The maps (Fig. 1) show that for England and Sweden the high excess mortality was spread across the countries. In England, the lowest excesses are in rural areas with lower population densities. The maps suggest the largest excess mortality in England were concentrated in the two largest cities, London and Birmingham especially for males. In Italy, the largest excesses in mortality were mostly in the north of the country in both urban and rural municipalities where the pandemic first hit and the impact was already significant before the lockdown in March 2020. Other areas of high excess include parts of Puglia and the islands of Sicily and Sardinia as well as large urban areas. In Sweden the maps show little or no regional clustering of excess mortality. The patterns visible for England and Italy are more pronounced for males than females.

**Fig. 1.**
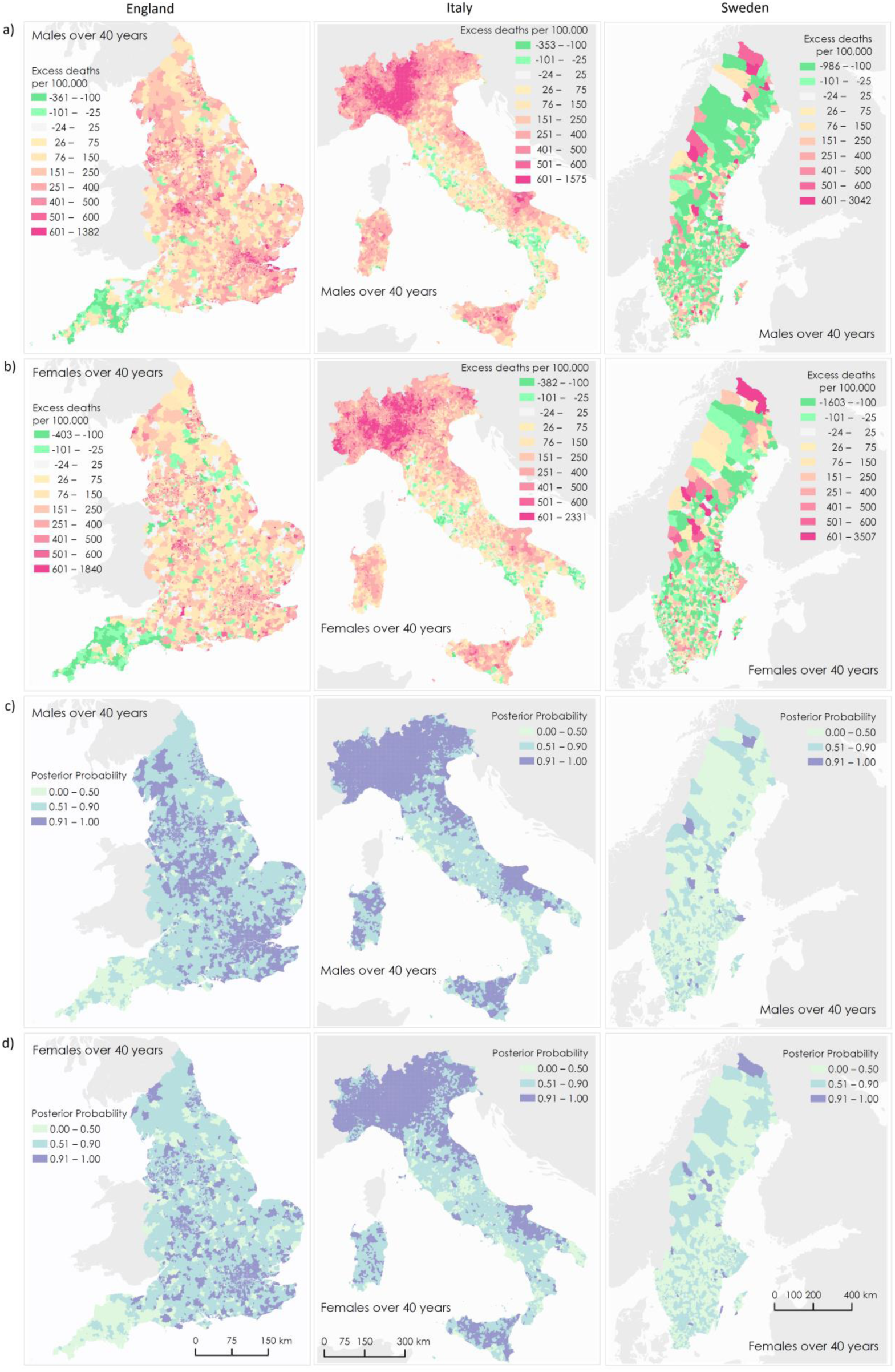
Maps of middle super output areas (MSOAs) in England (first column), Municipalities in Italy (second column) and DeSOs in Sweden (third column) showing excess deaths per 100,000 people aged 40 years and over. Excess deaths per 100,000 males (a) and females (b) from 1 March 2020 to 28 February 2021 compared to the same period for the preceding five years. Posterior probability that excess deaths > 0 for males (c) and females (d). The posterior probability measures the extent to which an estimate of excess/fewer deaths is likely to be a true increase/decrease. Where the entire posterior distribution of estimated excess deaths for an area is greater than zero, there is a posterior probability of ∼1 of a true increase, and conversely where the entire posterior distribution is less than zero there is a posterior probability of ∼0 of a true increase. This posterior probability would be ∼0.5 in an area in which an increase is statistically indistinguishable from a decrease. Contains OS data © Crown copyright (2020). Data available under the UK Open Government Licence v3.

### Associations of community characteristics with excess mortality

When considering the association between the covariates and excess mortality, there were striking similarities between the results for England and Sweden that were not always shared with Italy (Fig. 2). Considered individually (i.e. in univariate analysis), for England and Sweden, all covariates, except the workplace visits data showed positive associations with excess mortality across the categories for both sexes, although the association was generally weaker for females (Supplementary Tables 13, 15 & 17). For Italy, only PM_2.5_ and deprivation showed a positive association with excess mortality for both sexes. There were positive correlations between some of the covariates (Supplementary Tables 10-12); for example, Kendall’s Tau for England was 0.56 and 0.61 between overcrowding and NO_2_ and population density respectively, and for Sweden the equivalent correlations were 0.50 and 0.54. These contrast markedly with the same correlations for Italy (0.02 and 0.12). When the covariates were considered together (multivariate analysis), associations between excess deaths and NO_2_, and PM_2.5_ mostly or completely disappeared for England and Sweden. The association with population density and overcrowding/living area was attenuated, but remained, especially for males. For Italy the association with PM_2.5_ and deprivation remained in the multivariable analysis.

**Fig. 2.**
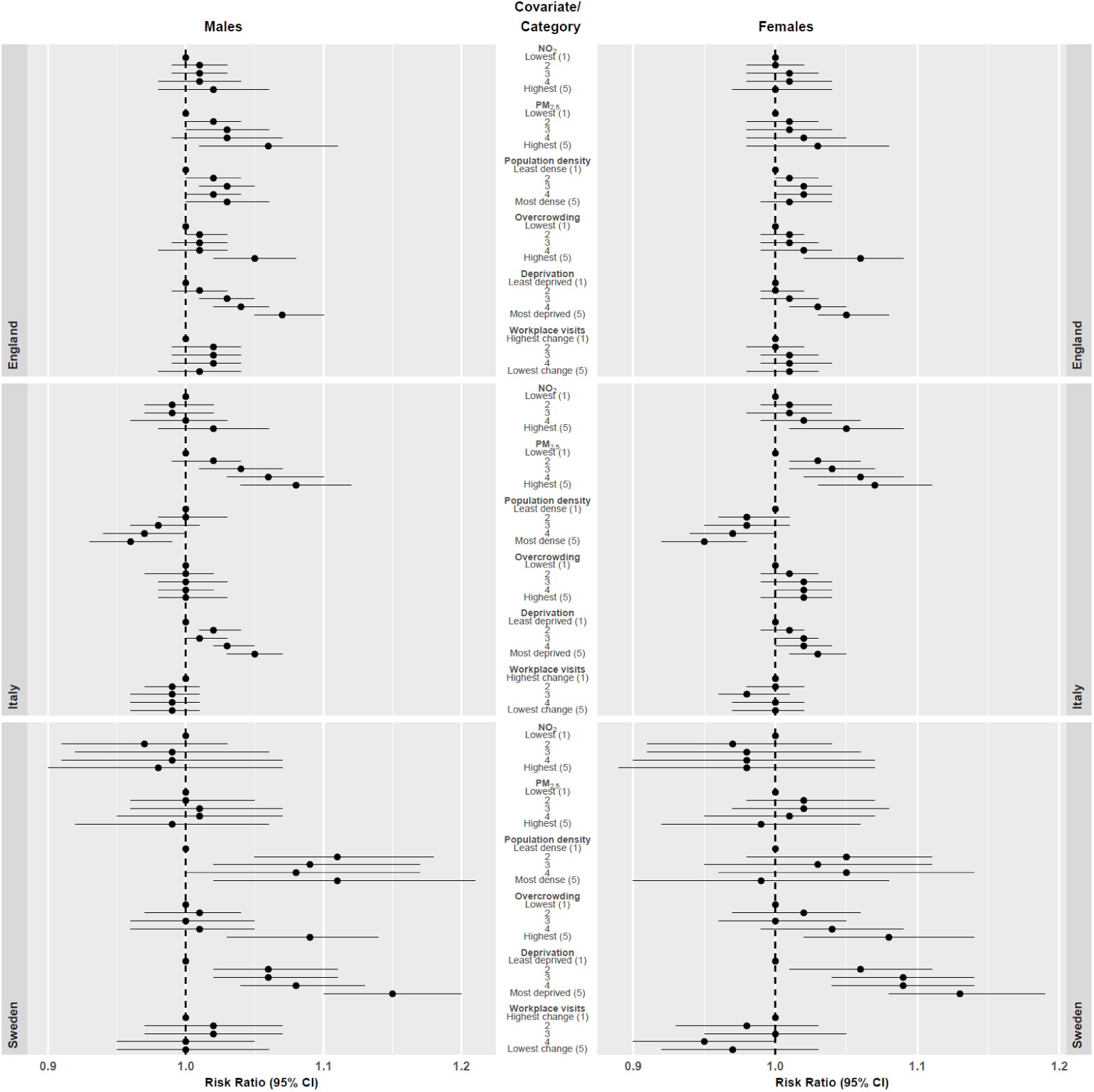
The relationship between community covariates of small areas in England, Italy and Sweden and excess mortality from 1 March 2020 to 28 Feb 2021 compared to the same period for the preceding five years. Proportional increase in death rates shown as rate ratios (data are presented as posterior mean with 95% credible intervals) for categories of the distributions relative to lowest category. Multivariable relationship between covariates and excess mortality after adjustment for the other covariates, numerical values reported in Supplementary Tables 14, 16 & 18.

Notably, the relationship with deprivation was strong under both univariate and multivariate analyses for all countries, both sexes. In particular, for Sweden there was a ∼15% (95% CI 10% - 20%) higher excess mortality for the most deprived category (compared to least deprived) for males and it was ∼13% (95% CI 8% - 19%) higher for females.

Aside from the positive association for deprivation, the results for Italy were distinct from the other two countries. Specifically: a weak negative association for both sexes with population density was present in the multivariable analysis for Italy whereas both England and Sweden showed a weak positive association; for both sexes, PM_2.5_ showed a strong positive association for Italy. This was much weaker for England, and non-existent for Sweden.

For all countries, the covariates accounted for a higher percentage of the variation in excess mortality across the community-level areas for males than females (Supplementary Tables 13-18).

### Results of sensitivity analyses

In sensitivity analysis, removing the random effect for local spatial correlation (Supplementary Tables 26-28) indicated a positive association, particularly for males for both air pollution covariates in the multivariable model and there also seemed to be an association for workplace visits for England. The results for Italy revealed a stronger association with NO_2_, and negative associations for population density and workplace visits for both sexes. The results for Sweden showed no material differences to the main model for this analysis. The sensitivity analyses using continuous variables for NO_2_, PM_2.5_ and population density (Supplementary Tables 20-25) showed no material differences to the main model, except a stronger association between PM_2.5_ and excess mortality for males in England (Supplementary Table 20).

## Discussion

Our study provides insights into the comparison of spatial distribution of excess mortality and the role of community-level characteristics in three European countries with dissimilar social structures which applied different public health strategies. As illustrated by Fig. 1, our maps revealed strong geographical patterns in England and Italy, but less so in Sweden. It supports our choices of geographical areas across the three countries to identify such patterns.

The similar patterns of results between England and Sweden suggest that the characteristics of communities affecting excess mortality in the first year of the COVID-19 pandemic are shared between some countries, in particular deprivation, even if the overall rates of excess mortality varied between countries. The contrasting results from Italy could be explained in at least two ways. Firstly the way the disease first spread within the country, specifically its rapid spread in the north in the initial phases before the national lockdown and clear response measures were in place, and a consequent high excess in mortality as shown in Fig. 1 and reported previously.^28^ This was not the case for England or Sweden where the disease spread later and more evenly.^29^ Secondly, because spatial units in Italy are based solely on administrative area, resulting in more diverse population sizes, with large metropolitan areas (such as Rome or Milan) aside small rural towns or villages. This also contributed to the high percentage of variance explained by local clustering (Table 3) for Italy. The heterogeneity in excess mortality observed within the cities in England and Sweden may have been present in large Italian cities, but was masked by the entire city comprising a single municipality.

Our results suggest a stronger deprivation gradient in excess mortality for Sweden than the other countries. Although we used different definitions of deprivation for the three countries and our data did not provide any direct explanations, previous research has shown a positive correlation between income and compliance with non-pharmaceutical interventions, such as handwashing, social distancing, reducing mobility and remote work.^30, 31^ Sweden’s reliance on recommendations rather than a strict lockdown^32^ could explain the stronger deprivation gradient, although more research is needed to confirm this hypothesis.

Sweden is a smaller country (2019 population 10.3 million) than both England (2019 population 56.3 million) and Italy (2019 population 59.7 million) with lower excess mortality per 100,000 population and DeSOs are smaller than both MSOAs and Italian municipalities. These differences implied wider 95% CIs around the estimates for Sweden than around those for England and Italy, for both the area-specific excess mortality estimates and the relationships between excess mortality and the covariates. The lower percentage of variance explained by local clustering (Table 3) and lack of regional patterns of excess mortality (Fig. 1) for Sweden could be explained by greater homogeneity across communities in the country.

This study not only provides insights into the community level factors affecting excess mortality, in particular the consistent association between deprivation and higher excess mortality shown for all three countries with different levels of overall deprivation, but also showed significant overlap in other important effects, for example, for both Sweden and England the multivariable results (Fig. 2) suggested that population density itself was a much less important driver of excess mortality than the univariable results at first suggested.

### Strengths and limitations

Our analysis aimed to use similar community-level areas and a range of covariates previously linked to excess mortality in COVID-19 studies across three European countries which experienced different pandemic dynamics and implemented different time-varying, public health interventions.^33^ The methodology applied combines a rigorous two-stage modelling approach, a large number areas, and expertise from each of the three countries.

The large range of population sizes for Italian municipalities is likely to affect the relationship between excess mortality and the covariates because the larger municipalities are unlikely to be homogenous communities with similar characteristics. The influence of the highly populous municipalities also affected the categories of the community characteristics. For example, 59.6% of the population over 40 years was in the top category of the population density covariate (Supplementary Table 19).

Some community characteristics previously reported to show associations with COVID-19 related excess mortality^5^ (proportion of population that is non-white, the number of care homes) could not be used here because the data (or close equivalents) were not available for all three countries. Although covariates were selected for consistency, it was not possible to use the same measures of deprivation or overcrowding for the three countries, meaning that they will be measuring different things. Furthermore, for the covariates which were the same or very similar (NO_2_, PM_2.5_, population density and workplace visits), the quintiles were defined per country, so cut-off points between quintiles were different for each country with Sweden having lower NO_2_ and PM_2.5_ quintile boundaries than England and Italy (Supplementary Tables 4-6) which should be considered when making direct comparisons between the results for the three countries for example in Fig. 2.

The process of estimating the counterfactual mortality rates for the community-level areas involves inherent uncertainty which might be reflected in some communities showing excess mortality even in the counterfactual case, so it was important that we accounted for all uncertainties in our model and reported results with 95% CIs. Consequently, our two stage model fully propagated uncertainty from the first stage (comparison period) to the second stage (study period).

We used mortality data for the comparison period to estimate the expected numbers of deaths in the study period. Prior to the start of the study period 29 COVID-19 related deaths had been recorded in Italy, and none in England and Sweden.^6^ Additional factors beyond the scope of this study, like temperature and healthcare provision are likely to have had an impact on excess mortality, both between the countries and between areas within countries. In addition, we used data for England from the last national census in 2011 to obtain information on sociodemographic characteristics of communities. To the extent that there have been demographic changes in the decade since then, this may have led to misclassification of areas with respect to their community characteristics.

The study period was chosen partly to avoid the effect of the vaccination programmes on COVID-19 mortality. By the end of the study period, approximately 2.1%, 4.0% and 6.5% of the populations over 40 years of England, Italy and Sweden had been given two vaccine doses, but the time delay between full vaccination and reaching maximum immunity combined with the likely time delay between infection and death meant that the vaccine programme will likely not have had a measurable effect on excess mortality by 28 Feb 2021.

We did not account for the spread or extent of infection in our model because seroprevalence data were not available at the high spatial resolution that we used. It is reasonable to assume that over the course of the year of the study period the disease had spread to all regions included in our study, although the higher excess mortality concentrated in the north of Italy strongly implies heterogeneity of infection rates in this country during the first wave of the pandemic.

Improving the comparability of community-level geographies and definitions of key covariates for epidemiological studies would greatly facilitate future international comparisons within Europe, allowing further research to build on our findings for future larger studies.

## Conclusion

In what we believe to be the first international study examining the role of community-level factors on excess mortality during the COVID-19 pandemic, we have shown striking similarities between England and Sweden in spite of the different government responses to the pandemic in the two countries as well as the very different population sizes and demographics and behaviours. Although the pandemic in Italy was unique, both in the timing and localized spread in the initial phases, some common aspects remain such as the role of deprivation. This strengthens the message to address deprivation related health inequalities before the next pandemic. In spite of the challenges inherent in making international comparisons with heterogeneous data, these results present a compelling reason to implement a cross country comparison in real time in future pandemics to help identify successful interventions. We believe the results presented here provide invaluable guidance for policy makers to target interventions such as localised public health messaging and provision of personal protective equipment that assist communities most at risk from mortality from this or future pandemics and that more interventions such as localised surge testing could have been implemented in high risk communities from an earlier stage.

## Data Availability

All data produced in the present study are available upon reasonable request to the authors

## Funding

The work of the UK Small Area Health Statistics Unit is overseen by UK Health Security Agency (UK HSA) and supported by the UK Medical Research Council, Grant number: MR/L01341X/1), and the National Institute for Health Research (NIHR) through its Health Protection Units (HPRUs) at Imperial College London in Environmental Exposures and Health and in Chemical and Radiation Threats and Hazards, and through Health Data Research UK (HDR UK). M.B., B.P., F.P., B.D. & D.F. acknowledge infrastructure support for the Department of Epidemiology and Biostatistics provided by the NIHR Imperial BRC. The views expressed are those of the authors. The work was partially supported by the MRC Centre for Environment and Health, which is funded by the Medical Research Council (MR/S019669/1, 2019-2024). This paper does not necessarily reflect the views of UK Health Security Agency, the National Institute for Health Research or the Department of Health and Social Care. C.B. and U.S. acknowledge funding from the Swedish Research Council for Health, Working-Life and Welfare under Grant 2020-00962 and the Swedish Cancer Society under Grant 20 0719.

## Acknowledgements

We thank Hima Daby, Gajanan Natu and Eric Johnson for their assistance in data acquisition, storage, preparation and governance; and the Office for National Statistics (www.ons.gov.uk) for the provision of mortality data derived from the national mortality registrations. M.S., F.D.D. and P.M. acknowledge the Italian National Institute of Statistics (ISTAT) for providing timely mortality data during the pandemic.

## Conflict of interest

The authors declare no conflicts of interest.

## Key-points

- Striking similarities in predictors of excess mortality between England and Sweden in spite of different government responses to the pandemic.
- Deprivation is a strong predictor of higher excess mortality in all three countries studies.
- These results provide useful guidance for policy makers to target public health interventions for this or future pandemics.

## Ethics and governance

### England

SAHSU holds approvals both from the London - South East Research Ethics Committee - reference 17/LO/0846 and from the Health Research Authority - Confidentiality Advisory Group - HRA CAG reference: 20/CAG/0028.

### Sweden

The European Regulation on the protection of natural persons with regard to the processing of personal data (GDPR) regulates the handling of personal information in Sweden. The Swedish mortality data were anonymized and delivered in aggregated form, which means that they do not constitute information on living persons as defined in GDPR. Therefore, their use is exempt from the requirement of ethical approval according to the Swedish Law of Research Ethics (2003:460).

### Italy

Italy is compliant with the European Regulation on data protection and security (GDPR) which regulates the handling of personal information in Italy. The Italian mortality data, made available online by the national Institute of Statistics (ISTAT) in aggregated form, thus is GDPR compliant and therefore is exempt for the requirement of ethics approval. Mortality data is collected under Regulation (EC) No 1338/2008 and N. 328/2011 of the European Parliament and of the Council on Community statistics on public health and health and safety at work, as regards statistics on causes of death.

## Data availability

### England

- SAHSU does not have permission to supply data to third parties. No identifiable information will be shared with any other organisation. Individual mortality data can be requested through the Office for National Statistics (https://www.ons.gov.uk/).
- Mid-year population estimates can be downloaded from https://www.ons.gov.uk/peoplepopulationandcommunity/populationandmigration/populationestimates/datasets/middlesuperoutputareamidyearpopulationestimates.
- English Index of Multiple Deprivation data can be downloaded from https://www.gov.uk/government/statistics/english-indices-of-deprivation-2019.
- 2011 Census data can be downloaded from https://www.ons.gov.uk/census/2011census/2011censusdata.
- Modelled air pollution data (NO_2_ & PM_2.5_) can be downloaded from https://ukair.defra.gov.uk/data/pcm-data.
- Workplace visits data provided by Google (https://www.google.com/covid19/mobility/).

### Italy

- Mortality, Population and living conditions data available from Istituto Nazionale di Statistica (ISTAT) (https://www.istat.it/it/archivio/240401).
- Italy Index of Multiple Deprivation data can be obtained upon request to the main authors of the referenced study ^23^.
- Italian air pollution data are not publicly available, but can be requested to the main authors of their publications ^15, 16^ upon request.
- Workplace visits data provided by Google (https://www.google.com/covid19/mobility/).

### Sweden

- Mortality and population data at DeSO level can be requested from Statistics Sweden (https://www.scb.se/en/).
- Deprivation covariate data are accessible from open-source ref ^12^.
- Data on overcrowding can be ordered from Statistics Sweden.
- Swedish air pollution data are not publicly available, but can be requested to the main authors of their publications ^15, 16^ upon request.
- Workplace visits data provided by Google (https://www.google.com/covid19/mobility/).

### Results

- Community level results (excess deaths, CIs, posterior probabilities) used in Fig. 1 are available on request to the authors.

## Code availability

The computer code written in R ^34^ for the two stages of Bayesian models used in this work is available on request to the authors.

## Notes

### Competing Interest Statement

The authors have declared no competing interest.

### Author Declarations

London - South East Research Ethics Committee - reference 17/LO/0846 gave ethical approval for this work Health Research Authority - Confidentiality Advisory Group - HRA CAG reference: 20/CAG/0028 gave ethical approval for this work

### Summary of Updates

Figs 1,2,3 merged into single figure, Fig. 1

