## Supplementary material for "Community factors and excess mortality in the COVID-19 pandemic in England, Italy and Sweden"

### Supplementary Section 1. Annual population estimates for England

Annual population was from the Office for National Statistics (ONS) mid-year (June) population estimates by age and sex MSOAs in England, 2015 to 2019. Mid-year population estimates for 2020 at MSOA level were unavailable at the time of writing, 2020 MSOA populations were estimated by distributing 2020 local authority district population estimates (released by ONS 25 June 2021)(1) according to MSOA shares in 2019. However, since the 2020 local authority district populations are mid-year estimates, the population denominators will be affected by the excess mortality during the first wave of the COVID-19 pandemic. To assess this effect we compared the ONS mid-year population estimates to a linear regression of the age stratified populations (2015-2019). The ONS mid-year population estimates take account of births and deaths, but also immigration, emigration and changes due to the demographics of the country(1) which is not the case for a simple linear regression. The population pyramid for England(2) shows that there are likely to be uniform changes in the 80+ age group, but non-uniform changes in the 40-59, 60-69, 70-79 age groups, and this is reflected in the comparison between the linear regression estimates against the ONS mid-year estimates (supplementary figure 1). The 80+ population age groups (M & F) linear regression estimates exceed the ONS mid-year estimates. We assert that this is due to the effect of the first wave of the pandemic where the majority of the COVID-19 deaths were in the 80+ age group(3). Consequently, for our 2020 population denominators we used the ONS mid-year estimates for the 40-59, 60-69, 70-79 age groups, and the linear regression estimates for the 80+ age group.

This issue is of little concern for Sweden, however: In 2020, considering the Swedish population aged 80+, there was < 5,00 excess deaths in relation to the reference period (in total, 58,906 persons aged 80+ died in 2020), which implies that <1% of the total 80+ denominator (80+ population size in 2020: ~540,000) could add to the overall error of the "pandemic year"-denominators (this error should otherwise be analogous to the errors in the denominators for each reference year). Hence, no adjustments of the denominator data were made for Sweden.

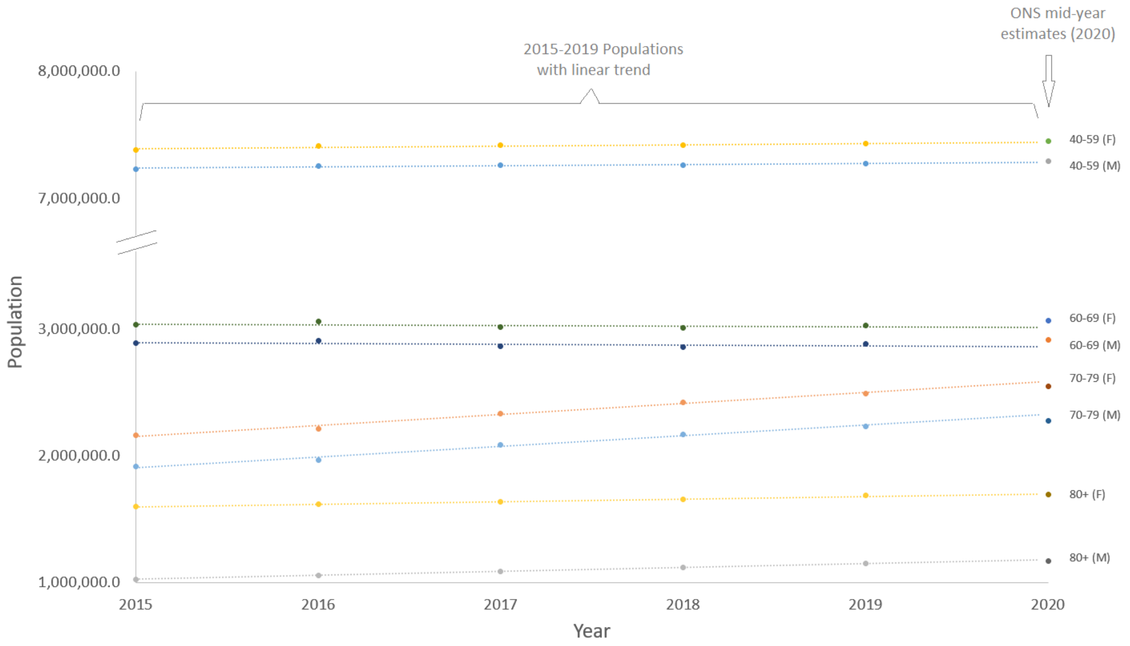


**Supplementary Fig. 1.** Population trends and estimates for England stratified by age and sex showing the comparison between 2015-2019 linear regression and the ONS 2020 mid-year population estimates.

**Supplementary Section 2. Details of statistical methods**

For the first stage model, we assumed that the number of deaths $y_{itk}$ for the $i$-th area ($i$=1,…6,791 for England; 1,…7,900 for Italy; 1,…5,985 for Sweden), the $t$-th year ($t$=2015,…,2019) of the comparison period and $k$-th age group ($k$=40-59, 60-69, 70-79, 80+) is modelled as a Poisson distribution:

$$y_{itk} \sim Poisson(\lambda_{ikt1} Pop_{itk})$$

where the log-transformed death rates are modelled as fixed and random effects:

$$log(\lambda_{ikt1}) = \alpha_0 + \beta_{0k} + b_{0i} + \gamma_t. \quad (1)$$

In (1) $\alpha_0$ is the common intercept which represents the overall mortality rate, while $\beta_{0k}$ is the age fixed effect for the *k-th* age group. To account for the multiple years in the comparison group, $\gamma_t$ provides a year-specific Gaussian random effect. Area-level effect $\boldsymbol{b}_0$ are modelled as a combination of a conditional autoregressive structure ($u_{0i}$), to capture local dependency and independent and identically distributed Gaussian random effects ($v_{0i}$) to account for the similarities of all the areas in the study region (i.e. each country):

$$b_{0i} = \frac{1}{\sqrt{\tau_b}}(\sqrt{1-\varphi}v_{0i} + \sqrt{\varphi}u_{0i}).$$

The model has a precision parameter ($\tau_b$), while $\varphi$ represents the weight of the spatially structured component (4). This model provides both local and global smoothing on the underlying death rate $\lambda_{ikt1}$.

We obtained the posterior distributions of the death rates in each area, age group and year, $\lambda_{ikt1}$; we then averaged over the 5 years of the comparison period to obtain $\lambda_{ik1}$, the expected death rate for the *i-th* area and *k-th* age group during the study period under the alternative scenario that the pandemic did not take place.

In the second stage model we estimated the ratio between death rates in the study period and the death rates we would have expected had there been no pandemic, using data for the comparison period. For the number of deaths in the $i$-th area and $k$-th age group in the study period, we specified the following model:

$$y_{i.2020.k} \sim Poisson(\rho_{ik}.\lambda_{ik1}.Pop_{i.2020.k})$$

where $\rho_{ik}$ represents the age-specific ratio between death rates in the study period and the comparison period ($\lambda_{ik1}$).

We modelled the ratio $\rho_{ik}$ in a similar way to stage one using terms to account for both space and age:

$$log(\rho_{ik}) = \alpha_1 + \beta_{1k} + \mathrm{b}_{1\mathrm{i}}. \tag{2}$$

Covariates were incorporated into this second stage log-linear model to evaluate their effect on the mortality rate ratio. For univariable effects in (2) we added the term $\delta X_i$ where $X_i$ is the category of the variable in the $i$-th area and $\delta$ is the associated effect. Similarly for the full multivariable model evaluating the joint effect of all variables we added $\sum_{\mathrm{n}=1}^{6}\sum_{\mathrm{j}=2}^{5}\delta_{\mathrm{nj}}\mathrm{X}_{\mathrm{inj}}$ with n=1,...6 covariates and $j$=2,…5 categories, as the first category is the reference one.

We obtained the posterior distribution of the estimated number of deaths across ages for the study period, calculated as $\hat{y}_{i.2020\cdot} = \sum_k \lambda_{ik1} \times \rho_{ik} \times Pop_{i.2020.k}$, and subtracted the corresponding number for 2020 using the rates for 2015-2019, calculated as $\hat{y}_{i.2015-2019\cdot} = \sum_k \lambda_{ik1.2015-2019} \times Pop_{i.2020.k}$ where the samples for $\lambda_{ik1.2015-2019}$ are drawn in equal proportions from each of the 5 years of the comparison period allowing the uncertainty to be fully represented. This difference was then divided by the 2020 population over 40 years old in that area and multiplied by 100,000.

**Supplementary Section 3. Results**

**Supplementary Table 1. Characteristics of the 6,791 Middle Super Output Areas in England.**

| | **Mean** | **Median** | **Range** | **Inter-quartile range** |
|---|---|---|---|---|
| Population* | 8,288 | 7,985 | 2,224 – 26,513 | 6,831 – 9,320 |
| Population* (M) | 4,098 | 3,926 | 1,089 – 14,535 | 3,348 – 4,629 |
| Population* (F) | 4,191 | 4,052 | 1,135 – 11,978 | 3,468 – 4,722 |
| Population over 40* (M) | 1,994 | 1,926 | 410 – 4,459 | 1,660 – 2,277 |
| Population over 40* (F) | 2,155 | 2,090 | 286 – 4,794 | 1,788 – 2,155 |
| Area ($km^2$) | 19.2 | 3.04 | 0.294 – 1,128 | 1.69 – 10.5 |
| Baseline deaths (M), per 100,000 males over 40, March 1 2015 – 28 February 2020 | 1,783 | 1,748 | 201 – 4,476 | 1,462 – 2,063 |
| Baseline deaths (F), per 100,000 females over 40, March 1 2015 – 28 February 2020 | 1,679 | 1,611 | 105 – 5,757 | 1,276 – 2,028 |

* 2019 mid-year population estimates.(5)

M – male; F – female; km- kilometre.

**Supplementary Table 2. Characteristics of the 7,900 Municipalities in Italy.**

| | **Mean** | **Median** | **Range** | **Inter-quartile range** |
|---|---|---|---|---|
| Population | 7,548 | 2,450 | 30 – 2,808,293 | 1,005 – 6,292 |
| Population (M) | 3,676 | 1,213 | 16 – 1,328,672 | 500 – 3,091 |
| Population (F) | 3,872 | 1,230 | 13 – 1,479,621 | 504 – 3,183 |
| Population over 40 (M) (2019) | 2,149 | 727 | 13 – 778,085 | 315 – 1,813 |
| Population over 40 (F) (2019) | 2,420 | 779 | 8 – 949,186 | 334 – 1,971 |
| Area ($km^2$) | 38.2 | 22.5 | 0.104 – 1,286 | 11.47 – 44.55 |
| Baseline deaths (M), per 100,000 males over 40, March 1 2015 – 28 February 2020 | 2,018 | 1,921 | 0 – 7,803 | 1,619 – 2,298 |
| Baseline deaths (F), per 100,000 females over 40, March 1 2015 – 28 February 2020 | 1,988 | 1,867 | 0 – 10,959 | 1,547 – 2,284 |

M – male; F – female; km- kilometre.

**Supplementary Table 3. Characteristics of the 5,985 DeSOs in Sweden.**

| | Mean | Median | Range | Inter-quartile range |
|---|---|---|---|---|
| Population | 1,726 | 1,708 | 663 – 5,291 | 1,402 – 2,018 |
| Population (M) | 868 | 858 | 335 – 2,543 | 701 – 1,016 |
| Population (F) | 876 | 849 | 319 – 2,748 | 695 – 1,006 |
| Population over 40 (M) (2019) | 428 | 420 | 4 – 1,131 | 346 - 502 |
| Population over 40 (F) (2019) | 445 | 437 | 4 – 1,233 | 357 - 528 |
| Area ($km^2$) | | | | |
| Baseline deaths (M), per 100,000 males over 40, March 1 2015 – 28 February 2020 | 1,716 | 1,513 | 0 – 7,862 | 1,101 - 2,120 |
| Baseline deaths (F), per 100,000 females over 40, March 1 2015 – 28 February 2020 | 1,667 | 1,271 | 0 – 11,395 | 851 – 2,128 |

M – male; F – female; km- kilometre.

**Supplementary Table 4. Characteristics of the study and comparison populations in England.**

| | Category | Nb. of MSOAs | Comparison period (1-3-2015 – 29-2-2020) Mean deaths per year (%) | Study period (1-3-2020 – 28-2-2021) Total deaths (%) |
|---|---|---|---|---|
| **Total** | | | 483,447 | 583,255 |
| **Sex** | | | | |
| Male | | | 237,052 (49.0) | 294,634 (50.5) |
| Female | | | 246,395 (50.0) | 288,621 (49.5) |
| **Age** | | | | |
| 40-59 | | | 41,120 (8.51) | 47,359 (8.12) |
| 60-69 | | | 56,363 (11.7) | 65,007 (11.1) |
| 70-79 | | | 107,969 (22.3) | 134,355 (23.0) |
| 80+ | | | 277,995 (57.5) | 336,534 (57.7) |
| **Place of death** | | | | |
| Hospital | | | 222,140 (45.9) | 249,854 (42.8) |
| Care home | | | 109,691 (22.7) | 138,411 (23.7) |
| Home | | | 114,389 (23.7) | 159,124 (27.3) |
| Hospice | | | 27,785 (5.75) | 23,896 (4.10) |
| Other/elsewhere | | | 9,442 (1.95) | 11,969 (2.05) |
| **$NO_2$ (μg/m$^3$)** | | | | |
| ≥ 3.60 to < 9.52 | 1 | 1359 | 114,116 (23.6) | 131,170 (22.5) |
| ≥ 9.52 to < 12.3 | 2 | 1358 | 106,942 (22.1) | 127,253 (21.8) |
| ≥ 12.3 to < 14.9 | 3 | 1358 | 97,141 (20.1) | 116,965 (20.1) |
| ≥ 14.9 to < 18.7 | 4 | 1358 | 95,201 (19.7) | 115,941 (19.9) |
| ≥ 18.7 to < 47.5 | 5 | 1358 | 70,047 (14.5) | 91,926 (15.8) |
| **$PM_{2.5}$ (μg/m$^3$)** | | | | |
| ≥ 5.05 to < 7.76 | 1 | 1359 | 110,181 (22.8) | 127,163 (21.8) |
| ≥ 7.76 to < 8.91 | 2 | 1358 | 104,549 (21.6) | 124,651 (21.4) |
| ≥ 8.91 to < 9.88 | 3 | 1358 | 104,134 (21.5) | 124,337 (21.3) |
| ≥ 9.88 to < 10.9 | 4 | 1358 | 94,103 (19.5) | 115,048 (19.7) |
| ≥ 10.9 to < 14.4 | 5 | 1358 | 70,480 (14.6) | 92,056 (15.8) |
| **Population density (population per km$^2$)** | | | | |
| ≥ 5.65 to < 456 | 1 | 1359 | 106,152 (22.0) | 123,814 (21.2) |
| ≥ 456 to < 1,877 | 2 | 1358 | 108,376 (22.4) | 129,217 (22.2) |
| ≥ 1,877 to < 3,445 | 3 | 1358 | 101,018 (20.9) | 121,962 (20.9) |
| ≥ 3,445 to < 5,340 | 4 | 1358 | 96,500 (20.0) | 116,880 (20.0) |
| ≥ 5,340 to < 29,000 | 5 | 1358 | 71,401 (14.8) | 91,382 (15.7) |
| **Overcrowding (% of overcrowded homes)** | | | | |
| ≥ 0.334 to < 1.65 | 1 | 1359 | 105,353 (21.8) | 123,775 (21.2) |
| ≥ 1.65 to < 2.42 | 2 | 1358 | 106,712 (22.1) | 126,385 (21.7) |
| ≥ 2.42 to < 3.67 | 3 | 1358 | 105,477 (21.8) | 125,937 (21.6) |
| ≥ 3.67 to < 6.26 | 4 | 1358 | 95,508 (19.8) | 114,643 (20.0) |
| ≥ 6.26 to < 36.5 | 5 | 1358 | 70,397 (14.6) | 92,515 (15.9) |
| **Deprivation (Index of Multiple Deprivation)** | | | | |
| ≥ 2.15 to < 10.4 | 1 | 1359 | 94,581 (19.6) | 113,243 (19.4) |

| | | | | |
|---|---|---|---|---|
| ≥ 10.4 to < 15.2 | 2 | 1358 | 99,800 (20.6) | 118,793 (20.4) |
| ≥ 15.2 to < 21.8 | 3 | 1358 | 98,264 (20.3) | 117,621 (20.2) |
| ≥ 21.8 to < 31.9 | 4 | 1358 | 93,347 (19.3) | 113,567 (19.5) |
| ≥ 31.9 to < 86.9 | 5 | 1358 | 97,456 (20.2) | 120,031 (20.6) |
| **Workplace visits (% change in travel to workplace)** | | | | |
| ≥ -68.7 to < -44.5 | 1 | 1358 | 72,164 (14.9) | 90,238 (15.5) |
| ≥ -44.4 to < -40.7 | 2 | 1373 | 93,098 (19.3) | 113,129 (19.4) |
| ≥ -40.7 to < -37.9 | 3 | 1364 | 100,301 (20.7) | 121,437 (20.8) |
| ≥ -37.9 to < -35.2 | 4 | 1351 | 108,031 (22.3) | 129,649 (22.2) |
| ≥ -35.2 to < -25.3 | 5 | 1345 | 109,853 (22.7) | 128,802 (22.1) |

$NO_2$ – nitrogen dioxide; $PM_{2.5}$ – particulate matter ≤ 2.5 μm diameter.

**Supplementary Table 5. Characteristics of the study and comparison populations in Italy**

| | Category | Nb. of municipalities | Comparison period (1-3-2015 – 29-2-2020) Mean deaths per year (%) | Study period (1-3-2020 – 28-2-2021) Total deaths (%) |
|---|---|---|---|---|
| **Total** | | | 635,168 | 751,893 |
| **Sex** | | | | |
| Male | | | 303,162 (47.7) | 366,065 (48.7) |
| Female | | | 332,006 (52.3) | 385,828 (51.3) |
| **Age (years)** | | | | |
| 40–59 | | | 39,635 (6.2) | 41,895 (5.6) |
| 60–69 | | | 59,221 (9.3) | 66,295 (8.8) |
| 70–79 | | | 127,949 (20.1) | 149,531 (19.9) |
| 80+ | | | 408,363 (64.3) | 494,172 (65.7) |
| **$NO_2$ (µg/m$^3$)** | | | | |
| ≥ 2.75 to ≤ 6.59 | 1 | 1,581 | 29,774 (4.7) | 31,744 (4.2) |
| > 6.59 to ≤ 9.51 | 2 | 1,577 | 57,579 (9.1) | 63,593 (8.5) |
| > 9.51 to ≤ 14.19 | 3 | 1,581 | 101,943 (16.0) | 114,043 (15.2) |
| > 14.19 to ≤ 19.40 | 4 | 1,580 | 136,565 (21.5) | 160,091 (21.3) |
| > 19.40 to ≤ 43.26 | 5 | 1,581 | 309,307 (48.7) | 382,422 (50.9) |
| **$PM_{2.5}$ (µg/m$^3$)** | | | | |
| ≥ 4.93 to ≤ 9.70 | 1 | 1,580 | 32,084 (5.1) | 35,734 (4.8) |
| > 9.70 to ≤ 11.41 | 2 | 1,578 | 66,637 (10.5) | 72,990 (9.7) |
| > 11.41 to ≤ 13.42 | 3 | 1,581 | 178,603 (28.1) | 201,788 (26.8) |
| > 13.42 to ≤ 17.93 | 4 | 1,580 | 187,267 (29.5) | 219,226 (29.2) |
| > 17.93 to ≤ 25.25 | 5 | 1,581 | 170,578 (26.9) | 222,155 (29.5) |
| **Population density (population per km$^2$)** | | | | |
| ≥ 0.8 to ≤ 34.1 | 1 | 1,581 | 25,064 (3.9) | 27,382 (3.6) |
| > 34.1 to ≤ 74.7 | 2 | 1,577 | 45,607 (7.2) | 51,467 (6.8) |
| > 74.7 to ≤ 151.6 | 3 | 1,581 | 69,930 (11.0) | 80,433 (10.7) |
| > 151.6 to ≤ 362.0 | 4 | 1,580 | 132,740 (20.9) | 156,606 (20.8) |
| > 362.0 to ≤ 12,177.8 | 5 | 1,581 | 361,827 (57.0) | 436,005 (58.0) |
| **Overcrowding (% of overcrowded homes)** | | | | |
| ≥ 0.0 to ≤ 0.3 | 1 | 1,509 | 25,070 (3.9) | 29,783 (4.0) |
| > 0.3 to ≤ 0.7 | 2 | 1,503 | 87,153 (13.7) | 104,190 (13.9) |
| > 0.7 to ≤ 1.0 | 3 | 1,938 | 171,533 (27.0) | 206,478 (27.5) |
| > 1.0 to ≤ 1.6 | 4 | 1,400 | 146,736 (23.1) | 175,154 (23.3) |
| > 1.6 to ≤ 16.7 | 5 | 1,550 | 204,676 (32.2) | 236,288 (31.4) |
| **Deprivation (index of deprivation)** | | | | |
| 1: very low | 1 | 1,550 | 200,208 (31.5) | 234,881 (31.2) |
| 2: low | 2 | 1,579 | 126,411 (19.9) | 151,228 (20.1) |
| 3: medium | 3 | 1,610 | 125,571 (19.8) | 149,043 (19.8) |

| | | | | |
|---|---|---|---|---|
| 4: high | 4 | 1,592 | 108,999 (17.2) | 128,325 (17.1) |
| 5: very high | 5 | 1,569 | 73,980 (11.6) | 88,416 (11.8) |
| **Workplace visits (% change in travel to workplace)** | | | | |
| ≥ -40.4 to ≤ -33.1 | 1 | 1,620 | 191,979 (30.2) | 233,226 (31.0) |
| > -33.1 to ≤ -30.5 | 2 | 1,619 | 121,060 (19.1) | 148,088 (19.7) |
| > -30.5 to ≤ -29.0 | 3 | 1,541 | 117,243 (18.5) | 136,376 (18.1) |
| > -29.0 to ≤ -27.6 | 4 | 1,554 | 85,023 (13.4) | 96,997 (12.9) |
| > -27.6 to ≤ -19.2 | 5 | 1,566 | 119,863 (18.9) | 137,206 (18.2) |

$NO_2$ – nitrogen dioxide; $PM_{2.5}$ – particulate matter ≤ 2.5 μm diameter.

**Supplementary Table 6. Characteristics of the study and comparison populations (Sweden)**

| | Category | Nb. of DeSOs | Comparison period (1-3-2015 – 29-2-2020) Mean deaths per year (%) | Study period (1-3-2020 – 28-2-2021) Total deaths (%) |
|---|---|---|---|---|
| **Total** | | | 88,709 | 97,997 |
| **Sex** | | | | |
| Male | | | 43,229 (48.7) | 49,207 (50.2) |
| Female | | | 45,480 (51.3) | 48,790 (49.8) |
| **Age (years)** | | | | |
| 40–59 | | | 4,832 (5.45) | 4,784 (4.88) |
| 60–69 | | | 9,042 (10.2) | 8,836 (9.02) |
| 70–79 | | | 19,873 (22.4) | 22,954 (23.4) |
| 80+ | | | 54,963 (62.0) | 61,423 (62.7) |
| **$NO_2$ (µg/m$^3$)** | | | | |
| ≥ 1.38 to ≤ 5.2 | 1 | 1,197 | 15,602 (17.6) | 15,507 (15.8) |
| > 5.2 to ≤ 8.51 | 2 | 1,197 | 18,617 (21.0) | 20,852 (21.3) |
| > 8.51 to ≤ 12.7 | 3 | 1,197 | 18,864 (21.3) | 22,108 (22.6) |
| > 12.7 to ≤ 16.3 | 4 | 1,197 | 18,571 (20.9) | 21,675 (22.1) |
| > 16.3 to ≤ 39.8 | 5 | 1,197 | 17,054 (19.2) | 17,855 (18.2) |
| **$PM_{2.5}$ (µg/m$^3$)** | | | | |
| ≥ 4.06 to ≤ 7.04 | 1 | 1,197 | 18,001 (20.3) | 19,201 (19.6) |
| > 7.04 to ≤ 7.66 | 2 | 1,197 | 17,960 (20.2) | 20,021 (20.4) |
| > 7.66 to ≤ 8.14 | 3 | 1,198 | 18,112 (20.4) | 20,326 (20.7) |
| > 8.14 to ≤ 8.8 | 4 | 1,196 | 17,019 (19.2) | 18,974 (19.4) |
| > 8.8 to ≤ 11.8 | 5 | 1,197 | 17,616 (19.9) | 19,475 (19.9) |
| **Population density (population per km$^2$)** | | | | |
| ≥ 0.0682 to ≤ 94.2 | 1 | 1,197 | 14,526 (16.4) | 15,507 (15.8) |
| > 94.2 to ≤ 383 | 2 | 1,197 | 18,923 (21.3) | 20,852 (21.3) |
| > 383 to ≤ 1,470 | 3 | 1,197 | 19,819 (22.3) | 21,675 (22.1) |
| > 1,470 to ≤ 4,850 | 4 | 1,197 | 19,398 (21.9) | 22,108 (22.6) |
| > 4,850 to ≤ 57,500 | 5 | 1,197 | 16,042 (18.1) | 17,855 (18.2) |
| **Overcrowding (living area, m$^2$ per person)** | | | | |
| ≤ 65.6 to > 46.8 | 1 | 1178 | 17,070 (19.2) | 18,340 (18.7) |
| ≤ 46.8 to > 43.5 | 2 | 1214 | 19,909 (22.4) | 21,758 (22.2) |
| ≤ 43.5 to > 40.2 | 3 | 1177 | 19,133 (21.6) | 20,941 (21.4) |
| ≤ 40.2 to > 35.2 | 4 | 1204 | 17,699 (20.0) | 19,767 (20.2) |
| ≤ 35.2 to ≥ 6.3 | 5 | 1212 | 14,897 (16.8) | 17,191 (17.5) |
| **Deprivation (% of inhabitants with low economic standard)** | | | | |
| ≥ 1.2 to ≤ 3.0 | 1 | 1,197 | 11,050 (12.5) | 12,768 (13.0) |
| > 13.0 to ≤ 18.9 | 2 | 1,197 | 15,045 (17.0) | 17,090 (17.4) |
| > 18.9 to ≤ 25.4 | 3 | 1,197 | 18,170 (20.5) | 19,847 (20.3) |
| > 25.4 to ≤ 34.4 | 4 | 1,197 | 21,718 (24.5) | 23,267 (23.7) |

| | | | | |
|---|---|---|---|---|
| > 34.4 to ≤ 94.8 | 5 | 1,197 | 22,726 (25.6) | 25,025 (25.5) |
| **Workplace visits (% change in travel to workplace)** | | | | |
| ≥ -48.3 to ≤ -30.7 | 1 | 1,198 | 14,386 (16.2) | 16,407 (16.7) |
| > -30.7 to ≤ -26.9 | 2 | 1,200 | 16,964 (19.1) | 18,841 (19.2) |
| > -26.9 to ≤ -24.9 | 3 | 1,196 | 17,190 (19.4) | 19,305 (19.7) |
| > -24.9 to ≤ -22.2 | 4 | 1,197 | 19,633 (22.1) | 21,074 (21.5) |
| > -22.2 to ≤ -12.3 | 5 | 1,194 | 20,535 (23.1) | 22,370 (22.8) |

$NO_2$ – nitrogen dioxide; $PM_{2.5}$ – particulate matter ≤ 2.5 µm diameter.

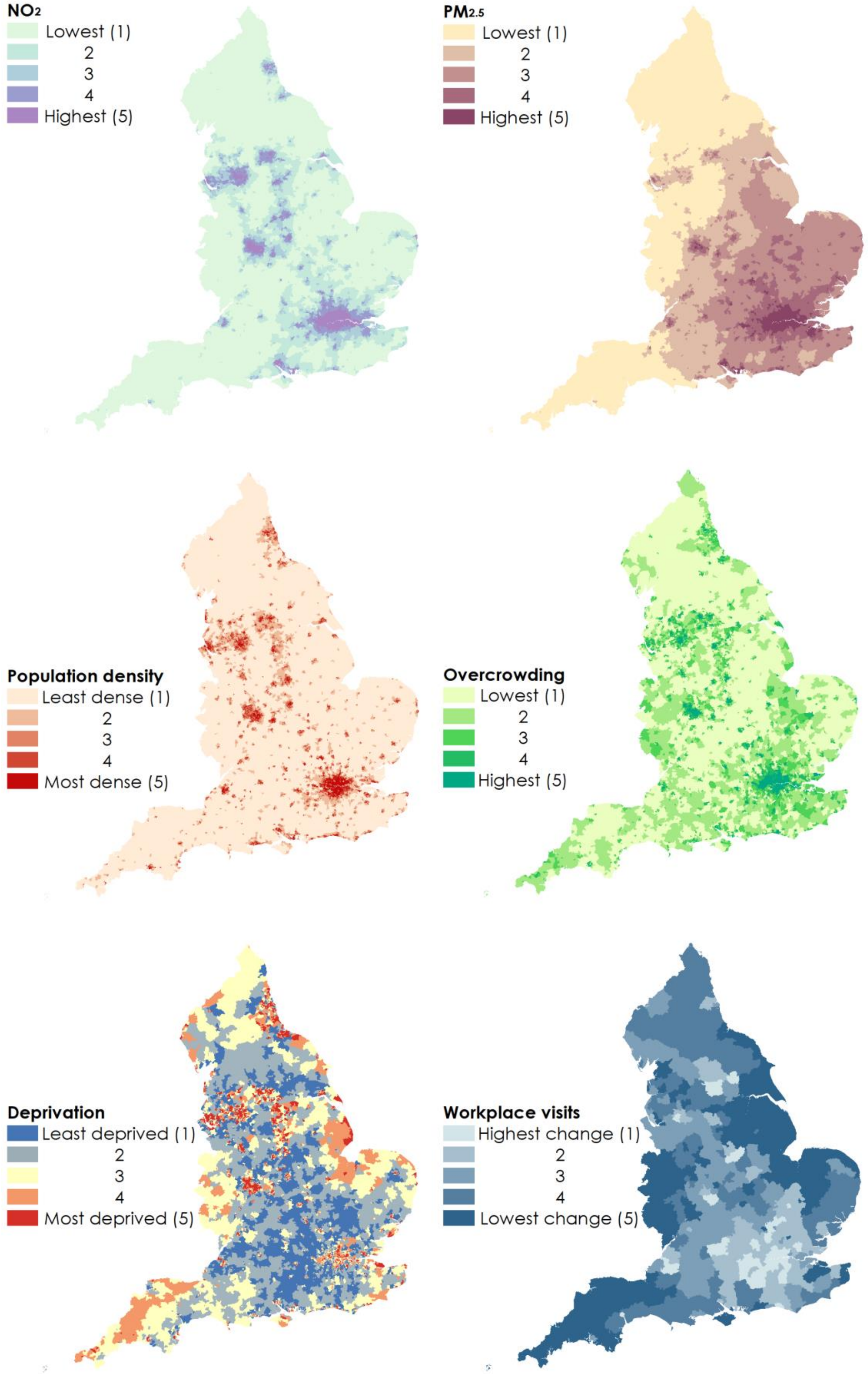


**Supplementary Fig. 2**. Choropleth maps of MSOAs in England showing categories of covariates. Note ‘workplace visits’ data were not available at MSOA resolution.

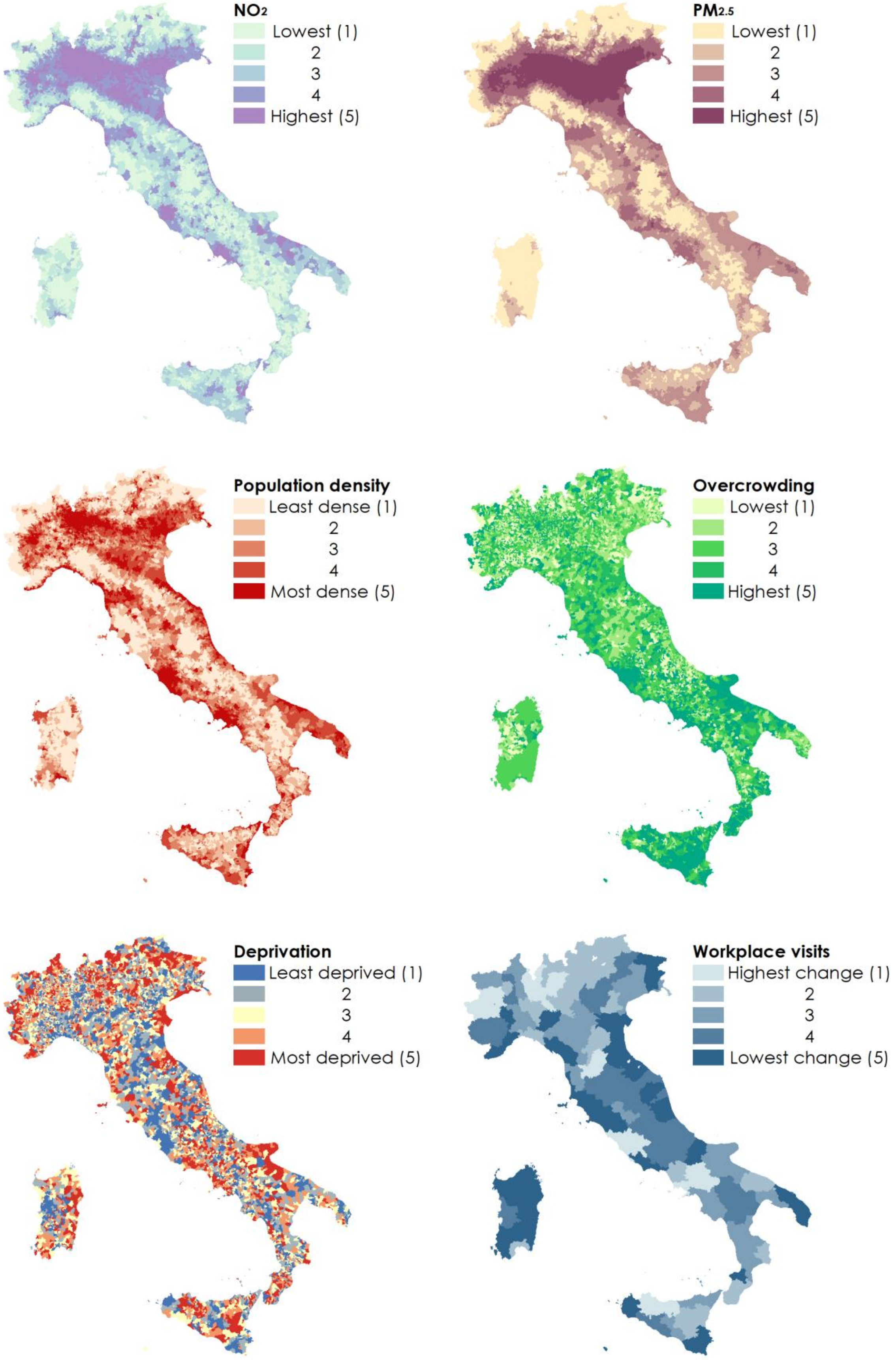


**Supplementary Fig. 3**. Choropleth maps of municipalities in Italy showing categories of covariates. Note 'workplace visits' data were not available at municipality resolution.

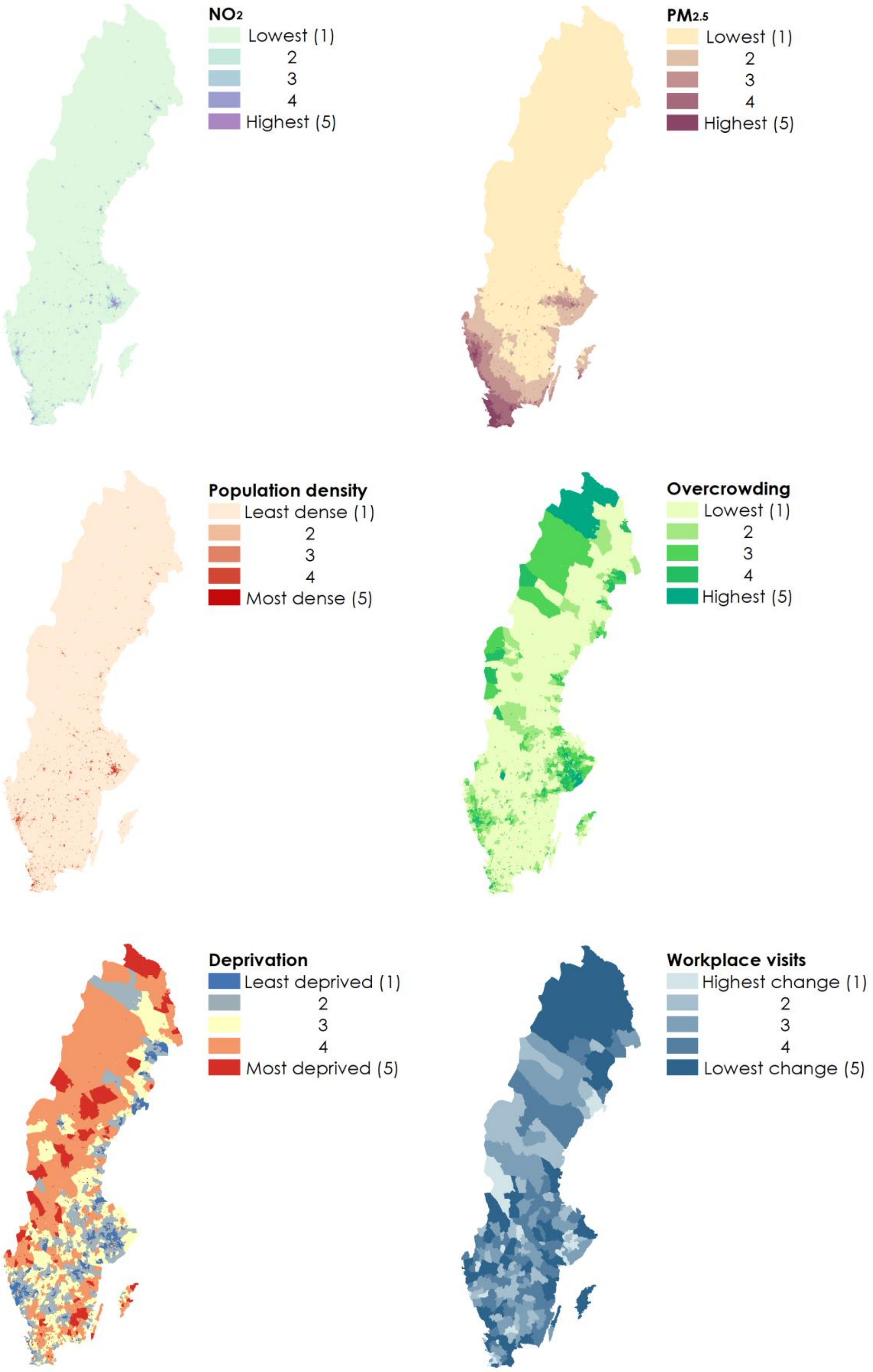


**Supplementary Fig. 4**. Choropleth maps of DeSOs in Sweden showing categories of covariates. Note 'workplace visits' data were not available at DeSO resolution.

**Supplementary Table 7. Excess death rates, percent increase in mortality and posterior probabilities for Middle Layer Super Output Areas in England.**

| | Mean | Median | Range | Inter-quartile range |
|---|---|---|---|---|
| Excess death rate (M), per 100,000 males over 40 | 316 | 291 | -362 – 1,383 | 173 - 441 |
| Excess death rate (F), per 100,000 females over 40 | 230 | 205 | -403 – 1,840 | 102 - 330 |
| Percent increase in mortality (M) | 17.8 | 16.5 | -16.3 – 82.9 | 9.90 – 24.3 |
| Percent increase in mortality (F) | 14.0 | 13.3 | -18.3 – 62.7 | 6.84 – 20.4 |
| Posterior probability that excess rate is greater than zero (M) | 0.881 | 0.946 | 0.024 – 1.00 | 0.834 – 0.990 |
| Posterior probability that excess rate is greater than zero (F) | 0.813 | 0.878 | 0.023 – 1.00 | 0.722 – 0.961 |

M – male; F – female.

**Supplementary Table 8. Excess death rates, percent increase in mortality and posterior probabilities for municipalities in Italy.**

| | Mean | Median | Range | Inter-quartile range |
|---|---|---|---|---|
| Excess death rate (M), per 100,000 males over 40 | 307 | 273 | -353 – 1,575 | 127 - 436 |
| Excess death rate (F), per 100,000 females over 40 | 278 | 249 | -382 – 2,332 | 108 - 402 |
| Percent increase in mortality (M) | 15.5 | 13.8 | -19.6 – 79.2 | 6.26 – 21.5 |
| Percent increase in mortality (F) | 14.0 | 13.0 | -24.3 – 59.6 | 5.66 – 20.8 |
| Posterior probability that excess rate is greater than zero (M) | 0.852 | 0.0.940 | 0.0007 – 1.00 | 0.761 – 0.993 |
| Posterior probability that excess rate is greater than zero (F) | 0.831 | 0.914 | 0.0000 – 1.00 | 0.729 – 0.987 |

M – male; F – female.

**Supplementary Table 9. Excess death rates, percent increase in mortality and posterior probabilities for DeSOs in Sweden.**

| | Mean | Median | Range | Inter-quartile range |
|---|---|---|---|---|
| Excess death rate (M), per 100,000 males over 40 | 156 | 94.9 | -986 – 3,042 | -44.1 – 278.8 |
| Excess death rate (F), per 100,000 females over 40 | 95.5 | 24.2 | -1,604 – 3,507 | -87.6 - 178 |
| Percent increase in mortality (M) | 9.19 | 7.07 | -28.8 - 102 | -2.24 – 18.3 |
| Percent increase in mortality (F) | 4.98 | 2.84 | -32.6 - 148 | -6.09 – 13.5 |
| Posterior probability that excess rate is greater than zero (M) | 0.588 | 0.592 | 0.0365 – 1.00 | 0.416 – 0.766 |
| Posterior probability that excess rate is greater than zero (F) | 0.518 | 0.504 | 0.0155 – 1.00 | 0.348 – 0.683 |

M – male; F – female.

**Supplementary Table 10. England: Correlation between community characteristics for Middle Super Output Areas in England:** Kendall's Tau coefficients between covariates (categories), n = 6,791 MSOAs.

| | $NO_2$ | $PM_{2.5}$ | Population density | Overcrowding | Deprivation | Workplace visits |
|---|---|---|---|---|---|---|
| **$NO_2$** | 1 | | | | | |
| **$PM_{2.5}$** | 0.55 | 1 | | | | |
| **Population density** | 0.59 | 0.39 | 1 | | | |
| **Overcrowding** | 0.56 | 0.42 | 0.61 | 1 | | |
| **Deprivation** | 0.24 | 0.01 | 0.33 | 0.51 | 1 | |
| **Workplace visits** | -0.41 | -0.42 | -0.30 | -0.26 | 0.14 | 1 |

**Supplementary Table 11. Italy: Correlation between community characteristics for Municipalities in Italy:** Kendall's Tau coefficients between covariates (categories), n = 7,900 municipalities.

| | $NO_2$ | $PM_{2.5}$ | Population density | Overcrowding | Deprivation | Workplace visits |
|---|---|---|---|---|---|---|
| **$NO_2$** | 1 | | | | | |
| **$PM_{2.5}$** | 0.77 | 1 | | | | |
| **Population density** | 0.63 | 0.55 | 1 | | | |
| **Overcrowding** | 0.02 | -0.03 | 0.12 | 1 | | |
| **Deprivation** | -0.11 | -0.11 | -0.13 | 0.15 | 1 | |
| **Workplace visits** | -0.30 | -0.23 | -0.25 | -0.07 | 0.00 | 1 |

**Supplementary Table 12. Sweden: Correlation between community characteristics for DeSOs in Sweden:** Kendall's Tau coefficients between covariates (categories), n = 5,985 DeSOs.

| | $NO_2$ | $PM_{2.5}$ | Population density | Overcrowding | Deprivation | Workplace visits |
|---|---|---|---|---|---|---|
| **$NO_2$** | 1 | | | | | |
| **$PM_{2.5}$** | 0.44 | 1 | | | | |
| **Population density** | 0.79 | 0.43 | 1 | | | |
| **Overcrowding** | 0.50 | 0.29 | 0.54 | 1 | | |
| **Deprivation** | 0.05 | 0.00 | 0.12 | 0.14 | 1 | |
| **Workplace visits** | -0.25 | -0.12 | -0.27 | -0.29 | 0.17 | 1 |

**Supplementary Table 13. England: One-variable-at-a-time models for MSOA (6,971) characteristics.** Age is included in all the models.

| Covariate | Category | Males | | Females | |
|---|---|---|---|---|---|
| | | Mean | 95% CI | Mean | 95% CI |
| **$NO_2$** | 1 | ref | ref | ref | ref |
| | 2 | 1.03 | 1.01 – 1.05 | 1.02 | 1.00 – 1.04 |
| | 3 | 1.06 | 1.04 – 1.07 | 1.04 | 1.02 – 1.06 |
| | 4 | 1.08 | 1.06 – 1.10 | 1.06 | 1.04 – 1.08 |
| | 5 | 1.13 | 1.10 – 1.16 | 1.09 | 1.07 – 1.12 |
| | % of variation explained by variable | 10.6 | 6.70 – 14.9 | 5.73 | 3.13 – 8.84 |
| **$PM_{2.5}$** | 1 | ref | ref | ref | ref |
| | 2 | 1.05 | 1.02 – 1.07 | 1.03 | 1.01 – 1.05 |
| | 3 | 1.08 | 1.06 – 1.11 | 1.05 | 1.02 – 1.08 |
| | 4 | 1.12 | 1.08 – 1.15 | 1.09 | 1.06 – 1.12 |
| | 5 | 1.18 | 1.14 – 1.22 | 1.13 | 1.09 – 1.17 |
| | % of variation explained by variable | 19.7 | 12.3 – 28.2 | 10.5 | 5.44 – 16.7 |
| **Population density** | 1 | ref | ref | ref | ref |
| | 2 | 1.04 | 1.02 – 1.05 | 1.03 | 1.01 – 1.04 |
| | 3 | 1.06 | 1.04 – 1.08 | 1.05 | 1.03 – 1.06 |
| | 4 | 1.06 | 1.05 – 1.08 | 1.06 | 1.04 – 1.08 |
| | 5 | 1.10 | 1.08 – 1.13 | 1.08 | 1.06 – 1.10 |
| | % of variation explained by variable | 6.49 | 4.28 – 9.03 | 4.29 | 2.53 – 6.43 |
| **Overcrowding** | 1 | ref | ref | ref | ref |
| | 2 | 1.03 | 1.01 – 1.05 | 1.02 | 1.00 – 1.03 |
| | 3 | 1.05 | 1.04 – 1.07 | 1.04 | 1.02 – 1.06 |
| | 4 | 1.07 | 1.05 – 1.09 | 1.06 | 1.05 – 1.08 |
| | 5 | 1.14 | 1.12 – 1.17 | 1.12 | 1.10 – 1.14 |
| | % of variation explained by variable | 11.9 | 8.70 – 15.4 | 8.87 | 6.11 – 12.0 |
| **Deprivation** | 1 | ref | ref | ref | ref |
| | 2 | 1.02 | 1.00 – 1.03 | 1.01 | 0.99 – 1.03 |
| | 3 | 1.05 | 1.03 – 1.06 | 1.03 | 1.01 – 1.04 |
| | 4 | 1.07 | 1.05 – 1.09 | 1.06 | 1.04 – 1.08 |
| | 5 | 1.11 | 1.10 – 1.13 | 1.10 | 1.08 – 1.12 |
| | % of variation explained by variable | 8.52 | 6.27 – 11.1 | 6.75 | 4.72 – 9.02 |
| **Workplace visits** | 1 | ref | ref | ref | ref |
| | 2 | 1.01 | 0.99 – 1.03 | 0.99 | 0.97 – 1.01 |
| | 3 | 1.00 | 0.98 – 1.03 | 1.00 | 0.97 – 1.02 |
| | 4 | 1.01 | 0.98 – 1.03 | 1.00 | 0.98 – 1.03 |
| | 5 | 1.01 | 0.98 – 1.04 | 1.01 | 0.98 – 1.03 |
| | % of variation explained by variable | 0.431 | 0.052 – 1.25 | 0.416 | 0.052 – 1.17 |

**Supplementary Table 14. England: Mutually adjusted multivariable models for all MSOA (6,971) characteristics.** Age is included in all the models.

| Covariate | Category | Males | | Females | |
|---|---|---|---|---|---|
| | | Mean | 95% CI | Mean | 95% CI |
| **$NO_2$** | 1 | ref | ref | ref | ref |
| | 2 | 1.01 | 0.99 – 1.03 | 1.00 | 0.98 – 1.02 |
| | 3 | 1.01 | 0.99 – 1.03 | 1.01 | 0.98 – 1.03 |
| | 4 | 1.01 | 0.98 – 1.04 | 1.01 | 0.98 – 1.04 |
| | 5 | 1.02 | 0.99 – 1.06 | 1.00 | 0.97 – 1.04 |
| **$PM_{2.5}$** | 1 | ref | ref | ref | ref |
| | 2 | 1.02 | 1.00 – 1.04 | 1.01 | 0.98 – 1.03 |
| | 3 | 1.03 | 1.00 – 1.06 | 1.01 | 0.98 – 1.04 |
| | 4 | 1.03 | 0.99 – 1.07 | 1.02 | 0.98 – 1.05 |
| | 5 | 1.06 | 1.01 – 1.11 | 1.03 | 0.98 – 1.08 |
| **Population density** | 1 | ref | ref | ref | ref |
| | 2 | 1.02 | 1.00 – 1.04 | 1.01 | 1.00 – 1.03 |
| | 3 | 1.03 | 1.01 – 1.05 | 1.02 | 1.00 – 1.04 |
| | 4 | 1.02 | 1.00 – 1.04 | 1.02 | 1.00 – 1.04 |
| | 5 | 1.03 | 1.00 – 1.06 | 1.01 | 0.99 – 1.04 |
| **Overcrowding** | 1 | ref | ref | ref | ref |
| | 2 | 1.01 | 1.00 – 1.03 | 1.01 | 0.99 – 1.02 |
| | 3 | 1.01 | 0.99 – 1.03 | 1.01 | 0.99 – 1.03 |
| | 4 | 1.01 | 0.98 – 1.03 | 1.02 | 0.99 – 1.04 |
| | 5 | 1.05 | 1.02 – 1.08 | 1.06 | 1.02 – 1.09 |
| **Deprivation** | 1 | ref | ref | ref | ref |
| | 2 | 1.01 | 0.99 – 1.03 | 1.00 | 0.99 – 1.02 |
| | 3 | 1.03 | 1.01 – 1.05 | 1.01 | 0.99 – 1.03 |
| | 4 | 1.04 | 1.02 – 1.06 | 1.03 | 1.01 – 1.05 |
| | 5 | 1.07 | 1.05 – 1.10 | 1.05 | 1.03 – 1.08 |
| **Workplace visits** | 1 | ref | ref | ref | ref |
| | 2 | 1.02 | 0.99 – 1.04 | 1.00 | 0.98 – 1.02 |
| | 3 | 1.02 | 0.99 – 1.04 | 1.01 | 0.99 – 1.03 |
| | 4 | 1.02 | 0.99 – 1.04 | 1.01 | 0.99 – 1.04 |
| | 5 | 1.01 | 0.98 – 1.04 | 1.01 | 0.98 – 1.03 |
| % of variation explained by all variables | | 17.7 | 13.1 – 23.0 | 11.2 | 8.07 – 14.8 |
| % of variance explained by local clustering for multivariable model | | 29.2 | 20.7 – 38.8 | 15.7 | 9.18 – 24.0 |
| % of variance explained by local clustering for model with no variables | | 45.4 | 36.6 – 54.5 | 27.1 | 19.6 – 35.7 |

**Total absolute excess across all MSOAs: M: 41,737; F: 32,827**

**Supplementary Table 15. Italy: One-variable-at-a-time models for Municipalities (7,900) characteristics.** Age is included in all the models.

| Covariate | Category | Males | | Females | |
|---|---|---|---|---|---|
| | | Mean | 95% CI | Mean | 95% CI |
| **$NO_2$** | 1 | ref | ref | ref | ref |
| | 2 | 1.00 | 0.98 – 1.02 | 1.02 | 1.00 – 1.04 |
| | 3 | 0.99 | 0.97 – 1.01 | 1.01 | 0.99 – 1.04 |
| | 4 | 1.01 | 0.98 – 1.02 | 1.03 | 1.00 – 1.05 |
| | 5 | 1.02 | 1.00 – 1.05 | 1.05 | 1.02 – 1.07 |
| | % of variation explained by variable | 1.07 | 0.36 – 2.09 | 1.74 | 0.52 – 3.51 |
| **$PM_{2.5}$** | 1 | ref | ref | ref | ref |
| | 2 | 1.00 | 0.98 – 1.02 | 1.02 | 1.00 – 1.05 |
| | 3 | 1.01 | 0.98 – 1.03 | 1.03 | 1.01 – 1.05 |
| | 4 | 1.03 | 1.01 – 1.06 | 1.05 | 1.02 – 1.07 |
| | 5 | 1.06 | 1.03 – 1.08 | 1.07 | 1.04 – 1.10 |
| | % of variation explained by variable | 3.20 | 1.21 – 5.85 | 3.33 | 1.20 – 6.22 |
| **Population density** | 1 | ref | ref | ref | ref |
| | 2 | 1.01 | 0.98 – 1.03 | 1.00 | 0.98 – 1.02 |
| | 3 | 1.00 | 0.97 – 1.02 | 1.01 | 0.99 – 1.03 |
| | 4 | 0.99 | 0.97 – 1.02 | 1.01 | 0.99 – 1.03 |
| | 5 | 0.99 | 0.97 – 1.02 | 1.00 | 0.98 – 1.03 |
| | % of variation explained by variable | 0.41 | 0.06 – 1.08 | 0.35 | 0.04 – 1.01 |
| **Overcrowding** | 1 | ref | ref | ref | ref |
| | 2 | 0.99 | 0.97 – 1.01 | 1.01 | 0.99 – 1.03 |
| | 3 | 1.00 | 0.98 – 1.02 | 1.02 | 1.00 – 1.04 |
| | 4 | 1.00 | 0.98 – 1.02 | 1.02 | 1.00 – 1.04 |
| | 5 | 1.00 | 0.98 – 1.03 | 1.02 | 1.00 – 1.04 |
| | % of variation explained by variable | 0.29 | 0.04 – 0.80 | 0.60 | 0.09 – 1.57 |
| **Deprivation** | 1 | ref | ref | ref | ref |
| | 2 | 1.02 | 1.01 – 1.04 | 1.01 | 1.01 – 1.03 |
| | 3 | 1.02 | 1.00 – 1.03 | 1.02 | 1.00 – 1.03 |
| | 4 | 1.03 | 1.02 – 1.05 | 1.02 | 1.01 – 1.04 |
| | 5 | 1.05 | 1.03 – 1.06 | 1.03 | 1.01 – 1.05 |
| | % of variation explained by variable | 1.52 | 0.66 – 2.61 | 0.78 | 0.22 – 1.63 |
| **Workplace visits** | 1 | ref | ref | ref | ref |
| | 2 | 0.99 | 0.96 – 1.01 | 1.00 | 0.97 – 1.02 |
| | 3 | 0.99 | 0.96 – 1.01 | 0.98 | 0.96 – 1.01 |
| | 4 | 0.98 | 0.96 – 1.01 | 0.99 | 0.96 – 1.02 |
| | 5 | 0.98 | 0.95 – 1.00 | 0.98 | 0.96 – 1.01 |
| | % of variation explained by variable | 0.73 | 0.11 – 1.87 | 0.71 | 0.09 – 1.85 |

**Supplementary Table 16. Italy: Mutually adjusted multivariable models for all Municipalities (7,900) characteristics.** Age is included in all the models.

| Covariate | Category | Males | | Females | |
|---|---|---|---|---|---|
| | | Mean | 95% CI | Mean | 95% CI |
| $NO_2$ | 1 | ref | ref | ref | ref |
| | 2 | 0.99 | 0.97 – 1.02 | 1.01 | 0.99 – 1.04 |
| | 3 | 0.99 | 0.97 – 1.02 | 1.01 | 0.98 – 1.04 |
| | 4 | 1.00 | 0.96 – 1.03 | 1.02 | 0.99 – 1.06 |
| | 5 | 1.02 | 0.98 – 1.06 | 1.05 | 1.01 – 1.09 |
| $PM_{2.5}$ | 1 | ref | ref | ref | ref |
| | 2 | 1.02 | 0.99 – 1.04 | 1.03 | 1.01 – 1.06 |
| | 3 | 1.04 | 1.01 – 1.07 | 1.04 | 1.01 – 1.07 |
| | 4 | 1.06 | 1.03 – 1.10 | 1.06 | 1.02 – 1.09 |
| | 5 | 1.08 | 1.04 – 1.12 | 1.07 | 1.03 – 1.11 |
| **Population density** | 1 | ref | ref | ref | ref |
| | 2 | 1.00 | 0.98 – 1.03 | 0.98 | 0.96 – 1.01 |
| | 3 | 0.98 | 0.96 – 1.01 | 0.98 | 0.95 – 1.01 |
| | 4 | 0.97 | 0.94 – 1.00 | 0.97 | 0.94 – 1.00 |
| | 5 | 0.96 | 0.93 – 0.99 | 0.95 | 0.92 – 0.98 |
| **Overcrowding** | 1 | ref | ref | ref | ref |
| | 2 | 1.00 | 0.97 – 1.02 | 1.01 | 0.99 – 1.03 |
| | 3 | 1.00 | 0.98 – 1.03 | 1.02 | 0.99 – 1.04 |
| | 4 | 1.00 | 0.98 – 1.02 | 1.02 | 1.00 – 1.04 |
| | 5 | 1.00 | 0.98 – 1.03 | 1.02 | 0.99 – 1.04 |
| **Deprivation** | 1 | ref | ref | ref | ref |
| | 2 | 1.02 | 1.01 – 1.04 | 1.01 | 0.99 – 1.02 |
| | 3 | 1.01 | 1.00 – 1.03 | 1.02 | 1.00 – 1.03 |
| | 4 | 1.03 | 1.02 – 1.05 | 1.02 | 1.00 – 1.04 |
| | 5 | 1.05 | 1.03 – 1.07 | 1.03 | 1.01 – 1.05 |
| **Workplace visits** | 1 | ref | ref | ref | ref |
| | 2 | 0.99 | 0.97 – 1.01 | 1.00 | 0.98 – 1.02 |
| | 3 | 0.99 | 0.96 – 1.01 | 0.98 | 0.96 – 1.01 |
| | 4 | 0.99 | 0.96 – 1.01 | 1.00 | 0.97 – 1.02 |
| | 5 | 0.99 | 0.96 – 1.01 | 1.00 | 0.97 – 1.02 |
| % of variation explained by all variables | | 7.74 | 4.67 – 11.7 | 6.44 | 3.57 – 10.3 |
| % of variance explained by local clustering for multivariable model | | 80.4 | 69.8 – 89.3 | 65.57 | 53.0 – 77.3 |
| % of variance explained by local clustering for model with no variables | | 81.1 | 71.3 – 89.5 | 65.12 | 52.8 – 76.3 |

**Total absolute excess across all municipalities: M: 43,122; F: 41,007**

**Supplementary Table 17. Sweden: One-variable-at-a-time models for DeSOs (5,985) characteristics.** Age is included in all the models.

| Covariate | Category | Males | | Females | |
|---|---|---|---|---|---|
| | | Mean | 95% CI | Mean | 95% CI |
| **$NO_2$** | 1 | ref | ref | ref | ref |
| | 2 | 1.05 | 1.01 – 1.09 | 1.01 | 0.97 – 1.05 |
| | 3 | 1.08 | 1.04 – 1.12 | 1.03 | 0.99 – 1.07 |
| | 4 | 1.10 | 1.06 – 1.15 | 1.04 | 1.00 – 1.08 |
| | 5 | 1.11 | 1.06 – 1.15 | 1.03 | 0.99 – 1.08 |
| | % of variation explained by variable | 2.69 | 1.16 – 4.61 | 0.57 | 0.086 – 1.44 |
| **$PM_{2.5}$** | 1 | ref | ref | ref | ref |
| | 2 | 1.04 | 1.00 – 1.09 | 1.04 | 1.00 – 1.09 |
| | 3 | 1.08 | 1.03 – 1.13 | 1.05 | 1.00 – 1.10 |
| | 4 | 1.08 | 1.02 – 1.13 | 1.03 | 0.98 – 1.08 |
| | 5 | 1.09 | 1.03 – 1.16 | 1.02 | 0.97 – 1.08 |
| | % of variation explained by variable | 1.98 | 0.45 – 4.28 | 0.72 | 0.12 – 1.78 |
| **Population density** | 1 | ref | ref | ref | ref |
| | 2 | 1.10 | 1.06 – 1.14 | 1.04 | 0.99 – 1.08 |
| | 3 | 1.09 | 1.05 – 1.13 | 1.03 | 0.99 – 1.07 |
| | 4 | 1.10 | 1.06 – 1.15 | 1.07 | 1.03 – 1.12 |
| | 5 | 1.18 | 1.12 – 1.23 | 1.04 | 0.99 – 1.09 |
| | % of variation explained by variable | 4.77 | 2.64 – 7.31 | 0.96 | 0.24 – 2.06 |
| **Overcrowding** | 1 | ref | ref | ref | ref |
| | 2 | 1.04 | 1.00 – 1.07 | 1.02 | 0.98 – 1.06 |
| | 3 | 1.05 | 1.01 – 1.09 | 1.01 | 0.98 – 1.06 |
| | 4 | 1.07 | 1.03 – 1.11 | 1.06 | 1.02 – 1.10 |
| | 5 | 1.20 | 1.15 – 1.25 | 1.12 | 1.07 – 1.17 |
| | % of variation explained by variable | 6.81 | 4.14 – 9.84 | 2.66 | 1.14 – 4.65 |
| **Deprivation** | 1 | ref | ref | ref | ref |
| | 2 | 1.07 | 1.02 – 1.11 | 1.06 | 1.01 – 1.10 |
| | 3 | 1.07 | 1.03 – 1.12 | 1.08 | 1.04 – 1.13 |
| | 4 | 1.10 | 1.06 – 1.15 | 1.09 | 1.04 – 1.14 |
| | 5 | 1.21 | 1.16 – 1.26 | 1.15 | 1.10 – 1.20 |
| | % of variation explained by variable | 6.62 | 4.24 – 9.38 | 3.34 | 1.64 – 5.39 |
| **Workplace visits** | 1 | ref | ref | ref | ref |
| | 2 | 1.03 | 0.98 – 1.09 | 1.00 | 0.95 – 1.05 |
| | 3 | 1.03 | 0.98 – 1.09 | 1.02 | 0.97 – 1.07 |
| | 4 | 1.02 | 0.96 – 1.07 | 0.97 | 0.92 – 1.02 |
| | 5 | 1.02 | 0.96 – 1.07 | 0.99 | 0.95 – 1.05 |
| | % of variation explained by variable | 0.662 | 0.0877 – 1.84 | 0.664 | 0.111 – 1.59 |

**Supplementary Table 18. Sweden: Mutually adjusted multivariable models for all DeSOs (5,985) characteristics.** Age is included in all the models.

| **Covariate** | **Category** | **Males** | | **Females** | |
|---|---|---|---|---|---|
| | | **Mean** | **95% CI** | **Mean** | **95% CI** |
| **$NO_2$** | 1 | ref | ref | ref | ref |
| | 2 | 0.97 | 0.91 – 1.03 | 0.97 | 0.91 – 1.04 |
| | 3 | 0.99 | 0.92 – 1.06 | 0.98 | 0.91 – 1.06 |
| | 4 | 0.99 | 0.91 – 1.07 | 0.98 | 0.90 – 1.07 |
| | 5 | 0.98 | 0.90 – 1.07 | 0.98 | 0.89 – 1.07 |
| **$PM_{2.5}$** | 1 | ref | ref | ref | ref |
| | 2 | 1.00 | 0.96 – 1.05 | 1.02 | 0.98 – 1.07 |
| | 3 | 1.01 | 0.96 – 1.07 | 1.02 | 0.97 – 1.08 |
| | 4 | 1.01 | 0.95 – 1.07 | 1.01 | 0.95 – 1.07 |
| | 5 | 0.99 | 0.92 – 1.06 | 0.99 | 0.92 – 1.06 |
| **Population density** | 1 | ref | ref | ref | ref |
| | 2 | 1.11 | 1.05 – 1.18 | 1.05 | 0.98 – 1.11 |
| | 3 | 1.09 | 1.02 – 1.17 | 1.03 | 0.95 – 1.11 |
| | 4 | 1.08 | 1.00 – 1.17 | 1.05 | 0.96 – 1.14 |
| | 5 | 1.11 | 1.02 – 1.21 | 0.99 | 0.90 – 1.08 |
| **Overcrowding** | 1 | ref | ref | ref | ref |
| | 2 | 1.01 | 0.97 – 1.04 | 1.02 | 0.97 - 1.06 |
| | 3 | 1.00 | 0.96 – 1.05 | 1.00 | 0.96 – 1.05 |
| | 4 | 1.01 | 0.96 – 1.05 | 1.04 | 0.99 – 1.09 |
| | 5 | 1.09 | 1.03 – 1.14 | 1.08 | 1.02 – 1.14 |
| **Deprivation** | 1 | ref | ref | ref | ref |
| | 2 | 1.06 | 1.02 – 1.11 | 1.06 | 1.01 – 1.11 |
| | 3 | 1.06 | 1.02 – 1.11 | 1.09 | 1.04 – 1.14 |
| | 4 | 1.08 | 1.04 – 1.13 | 1.09 | 1.04 – 1.14 |
| | 5 | 1.15 | 1.10 – 1.20 | 1.13 | 1.08 – 1.19 |
| **Workplace visits** | 1 | ref | ref | ref | ref |
| | 2 | 1.02 | 0.97 – 1.07 | 0.98 | 0.93 – 1.03 |
| | 3 | 1.02 | 0.97 – 1.07 | 1.00 | 0.95 – 1.05 |
| | 4 | 1.00 | 0.95 – 1.05 | 0.95 | 0.90 – 1.00 |
| | 5 | 1.00 | 0.95 – 1.06 | 0.97 | 0.92 – 1.03 |
| % of variation explained by all variables | | 11.3 | 8.23 – 14.8 | 6.44 | 4.24 – 9.08 |
| % of variance explained by local clustering for multivariable model | | 4.84 | 1.08 – 11.4 | 3.16 | 0.215 – 9.74 |
| % of variance explained by local clustering for model with no variables | | 8.45 | 3.24 – 16.2 | 3.74 | 0.444 – 10.1 |

**Total absolute excess across all DeSOs: M: 4,090; F: 2,987**

**Supplementary Table 19. Total excess mortality per 100,000 population stratified by covariates**

| England | | | | |
|---|---|---|---|---|
| **Covariate/Category** | **Males** | | **Females** | |
| | **Population > 40 years** | **Excess deaths per 100,000 (95% CI)** | **Population > 40 years** | **Excess deaths per 100,000 (95% CI)** |
| **$NO_2$** | | | | |
| 1 | 3,081,088 | 164 (92.3 – 236) | 3,374,801 | 125 (47.9 – 203) |
| 2 | 2,896,322 | 267 (196 – 339) | 3,172,105 | 196 (117 – 273) |
| 3 | 2,647,267 | 318 (248 – 390) | 2,885,370 | 239 (162 – 317) |
| 4 | 2,540,899 | 379 (309 – 452) | 2,736,587 | 293 (213 – 372) |
| 5 | 2,503,845 | 436 (378 – 495) | 2,592,859 | 289 (227 – 351) |
| **$PM_{2.5}$** | | | | |
| 1 | 2,840,956 | 204 (131 – 280) | 3,119,001 | 149 (67.9 – 230) |
| 2 | 2,745,513 | 298 (225 – 373) | 2,967,697 | 218 (138 – 298) |
| 3 | 2,894,494 | 297 (226 – 370) | 3,046,193 | 213 (134 – 291) |
| 4 | 2,695993 | 338 (272 – 408) | 2,914,299 | 264 (190 – 338) |
| 5 | 2,581,466 | 400 (344 – 457) | 2,714,530 | 277 (217 – 338) |
| **Population density** | | | | |
| 1 | 3,124,573 | 165 (97.3 – 233) | 3,378,801 | 120 (47.5 – 192) |
| 2 | 2,900,193 | 275 (204 – 348) | 3,189,299 | 201 (122 – 280) |
| 3 | 2,660,323 | 344 (273 – 418) | 2,918,606 | 248 (168 – 328) |
| 4 | 2,547,437 | 367 (296 – 441) | 2,763,657 | 289 (210 – 369) |
| 5 | 2,435,895 | 415 (355 – 476) | 2,511,358 | 284 (219 – 348) |
| **Overcrowding** | | | | |
| 1 | 3,084,486 | 174 (107 – 243) | 3,377,527 | 130 (57.1 – 203) |
| 2 | 2,946,650 | 243 (173 – 314) | 3,234,026 | 166 (90.2 – 243) |
| 3 | 2,728,326 | 313 (242 – 388) | 2,993,911 | 238 (156 – 319) |
| 4 | 2,469,095 | 373 (301 – 448) | 2,657,048 | 294 (214 – 375) |
| 5 | 2,439,866 | 469 (409 – 529) | 2,499,210 | 325 (262 – 389) |
| **Deprivation** | | | | |
| 1 | 2,974,471 | 200 (137 – 264) | 3,260,351 | 159 (90.4 – 228) |
| 2 | 2,930,299 | 223 (157 – 291) | 3,194,777 | 166 (93.5 – 239) |
| 3 | 2,787,724 | 288 (219 – 357) | 3,018,319 | 190 (115 – 265) |
| 4 | 2,595,358 | 359 (286 – 430) | 2,763,138 | 264 (188 – 341) |
| 5 | 2,380,568 | 500 (422 – 579) | 2,525,136 | 367 (283 – 452) |
| **Workplace visits** | | | | |
| 1 | 2,567,251 | 305 (248 – 363) | 2,721,171 | 229 (166 – 291) |
| 2 | 2,707,528 | 328 (261 – 396) | 2,926,776 | 232 (159 – 306) |
| 3 | 2,758,376 | 324 (254 – 396) | 2,999,571 | 232 (156 – 309) |
| 4 | 2,832,770 | 303 (231 – 378) | 3,077,028 | 228 (148 – 309) |
| 5 | 2,802,496 | 268 (194 – 346) | 3,037,175 | 191 (110 – 273) |

| Italy | | | | |
|---|---|---|---|---|
| **Covariate/Category** | **Males** | | **Females** | |
| | **Population > 40 years** | **Excess deaths per 100,000 (95% CI)** | **Population > 40 years** | **Excess deaths per 100,000 (95% CI)** |
| **$NO_2$** | | | | |
| 1 | 644,403 | 197 (102 – 325) | 687,282 | 155 (80.5 – 255) |
| 2 | 1,392,861 | 167 (85.6 – 280) | 1,512,831 | 156 (96.3 – 241) |
| 3 | 2,686,962 | 146 (74.5 – 247) | 2,967,178 | 135 (83.9 – 210) |
| 4 | 3,677,285 | 208 (140 – 306) | 4,110,085 | 200 (151 – 274) |
| 5 | 8,577,788 | 324 (262 – 415) | 9,840,544 | 257 (212 – 326) |
| **$PM_{2.5}$** | | | | |
| 1 | 763,545 | 214 (126 – 333) | 815,439 | 185 (117 – 277) |
| 2 | 1,628,935 | 137 (58.0 – 247) | 1,797,417 | 146 (89.5 – 229) |
| 3 | 4,673,230 | 167 (98.8 – 266) | 5,278,784 | 143 (95.8 – 217) |
| 4 | 5,037,521 | 222 (155 – 317) | 5,779,871 | 172 (125 – 244) |
| 5 | 4,876,068 | 413 (349 – 504) | 5,446,409 | 353 (306 – 425) |
| **Population density** | | | | |
| 1 | 513,480 | 272 (269 – 408) | 541,357 | 259 (177 – 366) |
| 2 | 1,045,298 | 266 (177 – 384) | 1,133,675 | 223 (159 – 314) |
| 3 | 1,761,388 | 224 (148 – 332) | 1,912,683 | 225 (168 – 307) |
| 4 | 3,655,269 | 218 (151 – 315) | 4,027,384 | 210 (162 – 283) |
| 5 | 10,003,864 | 268 (206 – 360) | 11,502,821 | 210 (166 – 279) |
| **Overcrowding** | | | | |
| 1 | 618,639 | 318 (229 – 437) | 662,835 | 281 (213 – 375) |
| 2 | 2,378,655 | 242 (171 – 342) | 2,619,250 | 222 (171 – 298) |
| 3 | 4,518,796 | 284 (216 – 382) | 5,072,981 | 252 (203 – 328) |
| 4 | 3,914,832 | 269 (201 – 365) | 4,415,126 | 239 (191 – 313) |
| 5 | 5,548,377 | 214 (149 – 309) | 6,347,728 | 155 (110 – 225) |
| **Deprivation** | | | | |
| 1 | 5,314,313 | 217 (151 – 313) | 6,180,280 | 183 (136 – 256) |
| 2 | 3,406,516 | 270 (201 – 368) | 3,816,519 | 229 (180 – 303) |
| 3 | 3,354,306 | 255 (186 – 354) | 3,733,344 | 230 (180 – 304) |
| 4 | 2,876,984 | 268 (198 – 368) | 3,187,766 | 219 (168 – 294) |
| 5 | 2,027,180 | 295 (223 – 395) | 2,200,011 | 240 (189 – 316) |
| **Workplace visits** | | | | |
| 1 | 5,384,911 | 295 (231 – 388) | 6,146,251 | 212 (167 – 282) |
| 2 | 3,295,644 | 308 (240 – 404) | 3,696,937 | 284 (235 – 358) |
| 3 | 3,100,402 | 234 (164 – 333) | 3,463,055 | 190 (140 – 265) |
| 4 | 2,165,927 | 177 (103 – 282) | 2,408,839 | 189 (135 – 269) |
| 5 | 3,032,415 | 192 (120 – 296) | 3,402,838 | 182 (130 – 260) |

| Sweden | | | | |
|---|---|---|---|---|
| **Covariate/Category** | **Males** | | **Females** | |
| | **Population > 40 years** | **Excess deaths per 100,000 (95% CI)** | **Population > 40 years** | **Excess deaths per 100,000 (95% CI)** |
| **$NO_2$** | | | | |
| 1 | 538,996 | 14.1 (-75.5 – 135) | 491,471 | 36.4 (-33.7 – 133) |
| 2 | 531,311 | 106 (14.3 – 234) | 542,043 | 67.1 (-13.5 – 182) |
| 3 | 540,865 | 167 (74.9 – 294) | 577,002 | 100 (22.3 – 210) |
| 4 | 496,679 | 222 (128 – 349) | 549,155 | 146 (65.7 – 263) |
| 5 | 476,938 | 235 (144 – 358) | 524,564 | 136 (58.1 – 248) |
| **$PM_{2.5}$ annual average (μg/m$^3$)** | | | | |
| 1 | 520,865 | 58.0 (-37.9 – 190) | 504,961 | 42.8 (-38.5 – 157) |
| 2 | 529,801 | 126 (35.6 – 253) | 540,039 | 112 (32.4 – 223) |
| 3 | 524,873 | 203 (113 – 326) | 556,214 | 133 (55.2 – 242) |
| 4 | 504,356 | 171 (81.6 – 292) | 538,166 | 103 (28.1 – 211) |
| 5 | 504,984 | 172 (81.4 – 295) | 544,855 | 96.1 (18.1 – 209) |
| **Population density** | | | | |
| 1 | 542,046 | -19.8 (-106 – 96.6) | 490,660 | 6.96 (-58.8 – 97.0) |
| 2 | 522,750 | 150 (56.1 – 280) | 530,260 | 93.0 (9.94 – 211) |
| 3 | 550,043 | 154 (61.1 – 282) | 590,957 | 88.6 (8.96 – 203) |
| 4 | 517,832 | 180 (85.0 – 309) | 569,707 | 176 (95.9 – 293) |
| 5 | 452,118 | 291 (202 – 412) | 502,651 | 116 (38.1 – 227) |
| **Overcrowding** | | | | |
| 1 | 534,722 | 27.2 (-65.0 – 155) | 517,249 | 20.7 (-52.7 – 124) |
| 2 | 545,288 | 98.5 (3.05 – 230) | 570,565 | 72.5 (-8.58 – 189) |
| 3 | 523,004 | 132 (38.2 – 262) | 559,185 | 60.1 (-20.2 – 176) |
| 4 | 512,233 | 158 (68.4 – 280) | 552,862 | 135 (57.7 – 246) |
| 5 | 469,542 | 338 (255 – 449) | 484,374 | 213 (137 – 319) |
| **Deprivation** | | | | |
| 1 | 546,658 | 23.7 (-42.4 – 114) | 546,019 | -6.61 (-59.0 – 66.6) |
| 2 | 541,944 | 95.9 (14.4 – 21.8) | 551,997 | 57.4 (-8.95 – 152) |
| 3 | 515,873 | 97.5 (4.26 – 227) | 538,973 | 95.9 (17.3 – 209) |
| 4 | 509,579 | 157 (50.7 – 304) | 541,417 | 112 (20.5 – 242) |
| 5 | 470,735 | 386 (273 – 542) | 505,829 | 244 (143 – 388) |
| **Workplace visits** | | | | |
| 1 | 512,245 | 167 (90.8 – 271) | 540,574 | 115 (47.6 – 209) |
| 2 | 517,072 | 158 (70.1 – 278) | 533,837 | 95.2 (20.1 – 203) |
| 3 | 504,867 | 174 (82.3 – 301) | 518,987 | 131 (52.0 – 242) |
| 4 | 513,474 | 113 (13.5 – 251) | 530,147 | 61.8 (-23.2 – 183) |
| 5 | 537,131 | 118 (17.7 – 257) | 560,690 | 89.2 (5.63 – 210) |

### Supplementary Section 4. Results of Sensitivity Analyses

### Sensitivity Analysis: Continuous Covariates

#### Supplementary Table 20. England: One-variable-at-a-time models.

| Variable | | Males | | Females | |
|---|---|---|---|---|---|
| | | Mean | 95% CI | Mean | 95% CI |
| **$NO_2$** | Linear effect (per IQR=7.27 μg/m$^3$) | 1.05 | 1.04 – 1.06 | 1.04 | 1.03 – 1.05 |
| **$PM_{2.5}$** | Linear effect (per IQR=2.51 μg/m$^3$) | 1.10 | 1.08 – 1.12 | 1.07 | 1.05 – 1.09 |
| **Population density** | Linear effect (per IQR=3,959 inhab./km$^2$) | 1.03 | 1.03 – 1.04 | 1.03 | 1.02 – 1.04 |

#### Supplementary Table 21. England: Mutually adjusted multivariable models. MALES

| Variable | | Males | | Females | |
|---|---|---|---|---|---|
| | | Mean | 95% CI | Mean | 95% CI |
| **$NO_2$** | Linear effect (per IQR=7.27 μg/m$^3$) | 9.98 | 0.96 – 1.00 | 1.00 | 0.98 – 1.02 |
| **$PM_{2.5}$** | Linear effect (per IQR=2.51 μg/m$^3$) | 1.06 | 1.03 – 1.09 | 1.02 | 0.99 – 1.05 |
| **Population density** | Linear effect (per IQR=3,959 inhab./km$^2$) | 1.00 | 0.99 – 1.01 | 1.00 | 0.99 – 1.01 |
| % of variance explained by local clustering for full model [a] | | 30.2 | 21.7 – 39.8 | 16.4 | 9.70 – 24.8 |

#### Supplementary Table 22. Italy: One-variable-at-a-time models.

| Variable | | Males | | Females | |
|---|---|---|---|---|---|
| | | Mean | 95% CI | Mean | 95% CI |
| **$NO_2$** | Linear effect (per IQR=10.8 μg/m$^3$) | 1.02 | 1.01 – 1.03 | 1.02 | 1.01 – 1.03 |
| **$PM_{2.5}$** | Linear effect (per IQR=6.84 μg/m$^3$) | 1.04 | 1.03 – 1.06 | 1.04 | 1.03 – 1.06 |
| **Population density** | Linear effect (per IQR=238 inhab./km$^2$) | 1.00 | 1.00 – 1.00 | 1.00 | 1.00 – 1.00 |

#### Supplementary Table 23. Italy: Mutually adjusted multivariable models.

| Variable | | Males | | Females | |
|---|---|---|---|---|---|
| | | Mean | 95% CI | Mean | 95% CI |
| **$NO_2$** | Linear effect (per IQR=10.8 μg/m$^3$) | 1.01 | 0.99 – 1.03 | 1.01 | 0.99 – 1.03 |
| **$PM_{2.5}$** | Linear effect (per IQR=6.84 μg/m$^3$) | 1.04 | 1.01 – 1.07 | 1.04 | 1.02 – 1.07 |
| **Population density** | Linear effect (per IQR=238 inhab./km$^2$) | 1.00 | 1.00 – 1.00 | 1.00 | 1.00 – 1.00 |
| % of variance explained by local clustering for full model [a] | | 80.7 | 70.0 – 89.6 | 66.4 | 53.7 – 78.1 |

#### Supplementary Table 24. Sweden: One-variable-at-a-time models.

| Variable | | Males | | Females | |
|---|---|---|---|---|---|
| | | Mean | 95% CI | Mea | 95% CI |
| **$NO_2$** | Linear effect (per IQR= 9.152 μg/m$^3$) | 1.05 | 1.03 – 1.07 | 1.01 | 0.98 – 1.03 |
| **$PM_{2.5}$** | Linear effect (per IQR= 1.401 μg/m$^3$) | 1.04 | 1.02 – 1.07 | 1.01 | 0.99 – 1.03 |
| **Population density** | Linear effect (per IQR= 3,384 inhab./km$^2$) | 1.01 | 1.01 – 1.02 | 1.00 | 0.99 – 1.01 |

#### Supplementary Table 25. Sweden: Mutually adjusted multivariable models. MALES

| Variable | | Males | | Females | |
|---|---|---|---|---|---|
| | | Mean | 95% CI | Mea | 95% CI |
| **$NO_2$** | Linear effect (per IQR= 9.152 μg/m$^3$) | 1.01 | 0.98 – 1.05 | 0.99 | 0.95 – 1.02 |
| **$PM_{2.5}$** | Linear effect (per IQR= 1.401 μg/m$^3$) | 1.01 | 0.98 – 1.05 | 1.01 | 0.98 – 1.04 |
| **Population density** | Linear effect (per IQR= 3,384 inhab./km$^2$) | 1.00 | 0.99 – 1.01 | 0.99 | 0.98 – 1.00 |
| % of variance explained by local clustering for full model [a] | | 4.97 | 1.21 – 11.58 | 3.08 | 0.262 – 8.99 |

[a] Model including age, $PM_{2.5}$ (linear), $NO_2$ (linear), deprivation index, population density (linear), overcrowding index and Google movement data

**Sensitivity Analysis: Models run without accounting for the local spatial correlation (no spatial smoothing)**

**Supplementary Table 26. England Multivariate model with no spatial smoothing (stage 1 & 2)**

| Variable | Category | Males | | Females | |
|---|---|---|---|---|---|
| | | **Mean** | **95% CI** | **Mean** | **95% CI** |
| **$NO_2$** | 1 | ref | ref | ref | ref |
| | 2 | 1.04 | 1.02 – 1.06 | 1.02 | 1.00 – 1.04 |
| | 3 | 1.05 | 1.03 – 1.07 | 1.03 | 1.01 – 1.05 |
| | 4 | 1.07 | 1.04 – 1.09 | 1.04 | 1.02 -1.06 |
| | 5 | 1.10 | 1.07 – 1.13 | 1.04 | 1.01 -1.07 |
| **$PM_{2.5}$** | 1 | ref | ref | ref | ref |
| | 2 | 1.02 | 1.01 – 1.04 | 1.02 | 1.00 – 1.03 |
| | 3 | 1.03 | 1.01 – 1.05 | 1.02 | 1.01 – 1.04 |
| | 4 | 1.03 | 1.02 – 1.05 | 1.04 | 1.02 – 1.06 |
| | 5 | 1.06 | 1.03 – 1.09 | 1.05 | 1.02 – 1.08 |
| **Population density** | 1 | ref | ref | ref | ref |
| | 2 | 1.02 | 1.00 – 1.03 | 1.01 | 0.99 – 1.03 |
| | 3 | 1.02 | 1.00 – 1.04 | 1.01 | 0.99 – 1.03 |
| | 4 | 1.01 | 0.99 – 1.03 | 1.01 | 0.99 – 1.03 |
| | 5 | 1.02 | 1.00 – 1.05 | 1.00 | 0.98 – 1.03 |
| **Overcrowded homes** | 1 | ref | ref | ref | ref |
| | 2 | 1.01 | 0.99 – 1.03 | 1.00 | 0.99 – 1.02 |
| | 3 | 1.00 | 0.99 – 1.02 | 1.01 | 0.99 – 1.03 |
| | 4 | 1.00 | 0.98 – 1.02 | 1.01 | 0.99 – 1.04 |
| | 5 | 1.05 | 1.02 – 1.08 | 1.05 | 1.02 – 1.09 |
| **IMD** | 1 | ref | ref | ref | ref |
| | 2 | 1.01 | 0.99 – 1.02 | 1.00 | 0.98 – 1.02 |
| | 3 | 1.02 | 1.01 – 1.04 | 1.00 | 0.99 – 1.02 |
| | 4 | 1.04 | 1.02 – 1.06 | 1.03 | 1.01 – 1.05 |
| | 5 | 1.07 | 1.05 – 1.10 | 1.06 | 1.03 – 1.09 |
| **Google movement (workplace)** | 1 | ref | ref | ref | ref |
| | 2 | 1.03 | 1.02 – 1.05 | 1.01 | 0.99 – 1.03 |
| | 3 | 1.05 | 1.03 – 1.07 | 1.03 | 1.01 – 1.05 |
| | 4 | 1.05 | 1.03 – 1.07 | 1.03 | 1.01 – 1.05 |
| | 5 | 1.03 | 1.01 – 1.05 | 1.01 | 0.99 – 1.03 |
| % of variation explained by all variables | | 27.7 | 24.0 – 31.5 | 16.2 | 13.2 – 19.2 |

**Total absolute excess across all MSOAs: M: 41,708 ; F: 32,801**

**Supplementary Table 27. Italy Multivariate model with no spatial smoothing (stage 1 & 2)**

| Variable | Category | Males | | Females | |
|---|---|---|---|---|---|
| | | **Mean** | **95% CI** | **Mean** | **95% CI** |
| **$NO_2$** | 1 | ref | ref | ref | ref |
| | 2 | 1.02 | 0.99 – 1.05 | 1.04 | 1.01 – 1.06 |
| | 3 | 1.04 | 1.01 – 1.07 | 1.06 | 1.03 – 1.09 |
| | 4 | 1.06 | 1.03 – 1.10 | 1.10 | 1.06 – 1.14 |
| | 5 | 1.10 | 1.06 – 1.15 | 1.13 | 1.09 – 1.17 |
| **$PM_{2.5}$** | 1 | ref | ref | ref | ref |
| | 2 | 0.99 | 0.97 – 1.08 | 1.00 | 0.97 – 1.03 |
| | 3 | 1.00 | 0.98 – 1.03 | 1.00 | 0.97 – 1.02 |
| | 4 | 1.04 | 1.00 – 1.07 | 1.03 | 0.99 – 1.06 |
| | 5 | 1.08 | 1.04 – 1.12 | 1.07 | 1.03 – 1.10 |
| **Population density** | 1 | ref | ref | ref | ref |
| | 2 | 0.99 | 0.96 – 1.02 | 0.97 | 0.95 – 1.00 |
| | 3 | 0.95 | 0.93 – 0.98 | 0.95 | 0.93 – 0.98 |
| | 4 | 0.92 | 0.90 – 0.95 | 0.92 | 0.90 – 0.95 |
| | 5 | 0.90 | 0.87 – 0.93 | 0.89 | 0.862 – 0.92 |
| **Household overcrowding index** | 1 | ref | ref | ref | ref |
| | 2 | 0.97 | 0.95 – 1.00 | 0.99 | 0.96 – 1.01 |
| | 3 | 0.98 | 0.96 – 1.00 | 1.00 | 0.98 – 1.02 |
| | 4 | 0.97 | 0.95 – 1.00 | 1.00 | 0.98 – 1.02 |
| | 5 | 0.96 | 0.93 – 0.98 | 0.98 | 0.95 – 1.00 |
| **Socio-economic status (Deprivation indicator)** | 1 | ref | ref | ref | ref |
| | 2 | 1.03 | 1.01 – 1.05 | 1.01 | 1.00 – 1.03 |
| | 3 | 1.02 | 1.00 – 1.04 | 1.02 | 1.00 – 1.04 |
| | 4 | 1.05 | 1.03 – 1.06 | 1.03 | 1.01 – 1.05 |
| | 5 | 1.07 | 1.05 – 1.09 | 1.04 | 1.02 – 1.06 |
| **Google movement (workplace)** | 1 | ref | ref | ref | ref |
| | 2 | 0.99 | 0.97 – 1.00 | 1.02 | 0.98 – 1.02 |
| | 3 | 0.94 | 0.92 – 0.96 | 0.95 | 0.96 – 0.97 |
| | 4 | 0.93 | 0.92 – 0.95 | 0.96 | 0.94 – 0.98 |
| | 5 | 0.95 | 0.93 – 0.96 | 0.96 | 0.95 – 0.98 |
| % of variation explained by all variables | | 19.2 | 16.02 – 22.6 | 14.8 | 11.9 – 17.9 |

**Total absolute excess across all municipalities: M: 43,361; F: 41,204**

**Supplementary Table 28. Sweden Multivariate model with no spatial smoothing (stage 1 & 2)**

| Variable | Category | Males | | Females | |
|---|---|---|---|---|---|
| | | **Mean** | **95% CI** | **Mean** | **95% CI** |
| **$NO_2$** | 1 | ref | ref | ref | ref |
| | 2 | 0.97 | 0.91 – 1.02 | 0.97 | 0.91 – 1.04 |
| | 3 | 0.99 | 0.92 – 1.06 | 0.98 | 0.91 – 1.06 |
| | 4 | 0.99 | 0.92 – 1.07 | 0.99 | 0.91 – 1.07 |
| | 5 | 0.99 | 0.91 – 1.07 | 0.98 | 0.90 – 1.07 |
| **$PM_{2.5}$** | 1 | ref | ref | ref | ref |
| | 2 | 1.00 | 0.96 – 1.04 | 1.02 | 0.98 – 1.06 |
| | 3 | 1.02 | 0.98 – 1.06 | 1.02 | 0.97 – 1.06 |
| | 4 | 1.00 | 0.96 – 1.05 | 1.01 | 0.96 – 1.05 |
| | 5 | 0.99 | 0.95 – 1.04 | 0.99 | 0.95 – 1.04 |
| **Population density** | 1 | ref | ref | ref | ref |
| | 2 | 1.11 | 1.05 – 1.18 | 1.04 | 0.98 – 1.11 |
| | 3 | 1.09 | 1.01 – 1.17 | 1.03 | 0.95 – 1.11 |
| | 4 | 1.08 | 1.00 – 1.16 | 1.05 | 0.96 – 1.14 |
| | 5 | 1.11 | 1.02 – 1.21 | 0.98 | 0.90 – 1.08 |
| **Living area** | 1 | ref | ref | ref | ref |
| | 2 | 1.00 | 0.96 – 1.04 | 1.02 | 0.97 – 1.06 |
| | 3 | 1.01 | 0.97 – 1.05 | 1.00 | 0.95 – 1.04 |
| | 4 | 1.01 | 0.97 – 1.06 | 1.04 | 0.99 – 1.09 |
| | 5 | 1.10 | 1.05 – 1.16 | 1.08 | 1.02 – 1.14 |
| **Economic status (Deprivation indicator SDI1)** | 1 | ref | ref | ref | ref |
| | 2 | 1.06 | 1.02 – 1.11 | 1.06 | 1.02 – 1.11 |
| | 3 | 1.06 | 1.02 – 1.10 | 1.09 | 1.04 – 1.14 |
| | 4 | 1.08 | 1.04 – 1.12 | 1.09 | 1.04 – 1.14 |
| | 5 | 1.14 | 1.09 – 1.19 | 1.14 | 1.09 – 1.19 |
| **Google movement (workplace)** | 1 | ref | ref | ref | ref |
| | 2 | 1.01 | 1.97 – 1.05 | 0.98 | 0.94 – 1.02 |
| | 3 | 1.01 | 0.97 – 1.06 | 1.00 | 0.95 – 1.04 |
| | 4 | 0.99 | 0.95 – 1.04 | 0.95 | 0.91 – 0.99 |
| | 5 | 0.99 | 0.95 – 1.04 | 0.98 | 0.93 – 1.02 |
| % of variation contributed by all variables | | 12.73 | 9.52 – 16.30 | 6.22 | 4.12 – 8.60 |

**Total absolute excess across all DeSOs: M: 3,727; F: 2,625**
